# A Quantitative Lung Computed Tomography Image Feature for Multi-Center Severity Assessment of COVID-19

**DOI:** 10.1101/2020.07.13.20152231

**Authors:** Biswajoy Ghosh, Nikhil Kumar, Nitisha Singh, Anup K. Sadhu, Nirmalya Ghosh, Pabitra Mitra, Jyotirmoy Chatterjee

## Abstract

The COVID-19 pandemic has affected millions and congested healthcare systems globally. Hence an objective severity assessment is crucial in making therapeutic decisions judiciously. Computed Tomography (CT)-scans can provide demarcating features to identify severity of pneumonia —commonly associated with COVID-19—in the affected lungs. Here, a quantitative severity assessing chest CT image feature is demonstrated for COVID-19 patients. We incorporated 509 CT images from 101 diagnosed and expert-annotated cases (age 20-90, 60% males) in the study collected from a multi-center Italian database^1^ sourced from 41 radio-diagnostic centers. Lesions in the form of opacifications, crazy-paving patterns, and consolidations were segmented. The severity determining feature —*L*_*norm*_ was quantified and established to be statistically distinct for the three —mild, moderate, and severe classes (p-value<0.0001). The thresholds of *L*_*norm*_ for a 3-class classification were determined based on the optimum sensitivity/specificity combination from Receiver Operating Characteristic (ROC) analyses. The feature *L*_*norm*_ classified the cases in the three severity categories with 86.88% accuracy. ‘Substantial’ to ‘almost-perfect’ intra-rater and inter-rater agreements were achieved involving expert (manual segmentation) and non-expert (graph-cut and deep-learning based segmentation) labels (*κ*-score 0.79-0.97). We trained several machine learning classification models and showed *L*_*norm*_ alone has a superior diagnostic accuracy over standard image intensity and texture features. Classification accuracy was further increased when *L*_*norm*_ was used for 2-class classification i.e. to delineate the severe cases from non-severe ones with a high sensitivity (97.7%), and specificity (97.49%). Therefore, key highlights of the COVID-19 severity assessment feature are high accuracy, low dependency on expert availability, and wide utility across different CT-imaging centers.

## 1 Introduction

With the onset of the COVID-19 pandemic caused by the SARS-CoV-2 coronavirus, newer tools and techniques are increasingly needed for efficient detection and therapy. As of now, RT-PCR based detection of the virus from oral/nasal swabs is globally accepted as the confirmatory test. However, due to the chances of the absence of viral particles on the swab especially in the asymptomatic or mild cases, the sensitivity of the method suffers (71% [1]). Therefore several screening methods are being deployed to augment COVID-19 detection including clinical history, symptom assessment, blood tests and imaging methods. Among the different imaging methods, chest X-Ray [2], Computed Tomography (CT) [3], and Ultrasonography (USG) [4, 5] are used in different clinical settings across the world to identify features associated with lung pneumonia commonly caused in the COVID-19 infection. Among these imaging methods, CT and high-resolution CT (HRCT) have shown a sensitivity of up to 98% [1] and hence has emerged as a strong screening tool for COVID-19 [3]. Thus, CT-imaging reduces the incidence of several infected individuals being discharged back in the community [6, 7].

The most common pathology seen in COVID-19 is pneumonia [7] (Figure 1) which eventually disrupts the lungs’ ability for gaseous exchange and reduces the oxygen availability for the normal cells to function. Due to the excessive deposition of fluid and pus (exudates) in the alveolar space, breathing is obstructed leading to respiratory failure. In severe cases, the immune response goes systemic and damages other vital organs like heart [8], liver [9] and kidney [10]. This increases the mortality of people with already present underlying health conditions and comorbidities like CAD (coronary artery disease), COPD (chronic obstructive pulmonary disease), CRF (chronic renal failure), diabetes, cancer, hepatitis, immunodeficiency etc [11]. Although the disease can be fatal, most individuals show only mild symptoms and do not need hospitalization.

**Figure 1:**
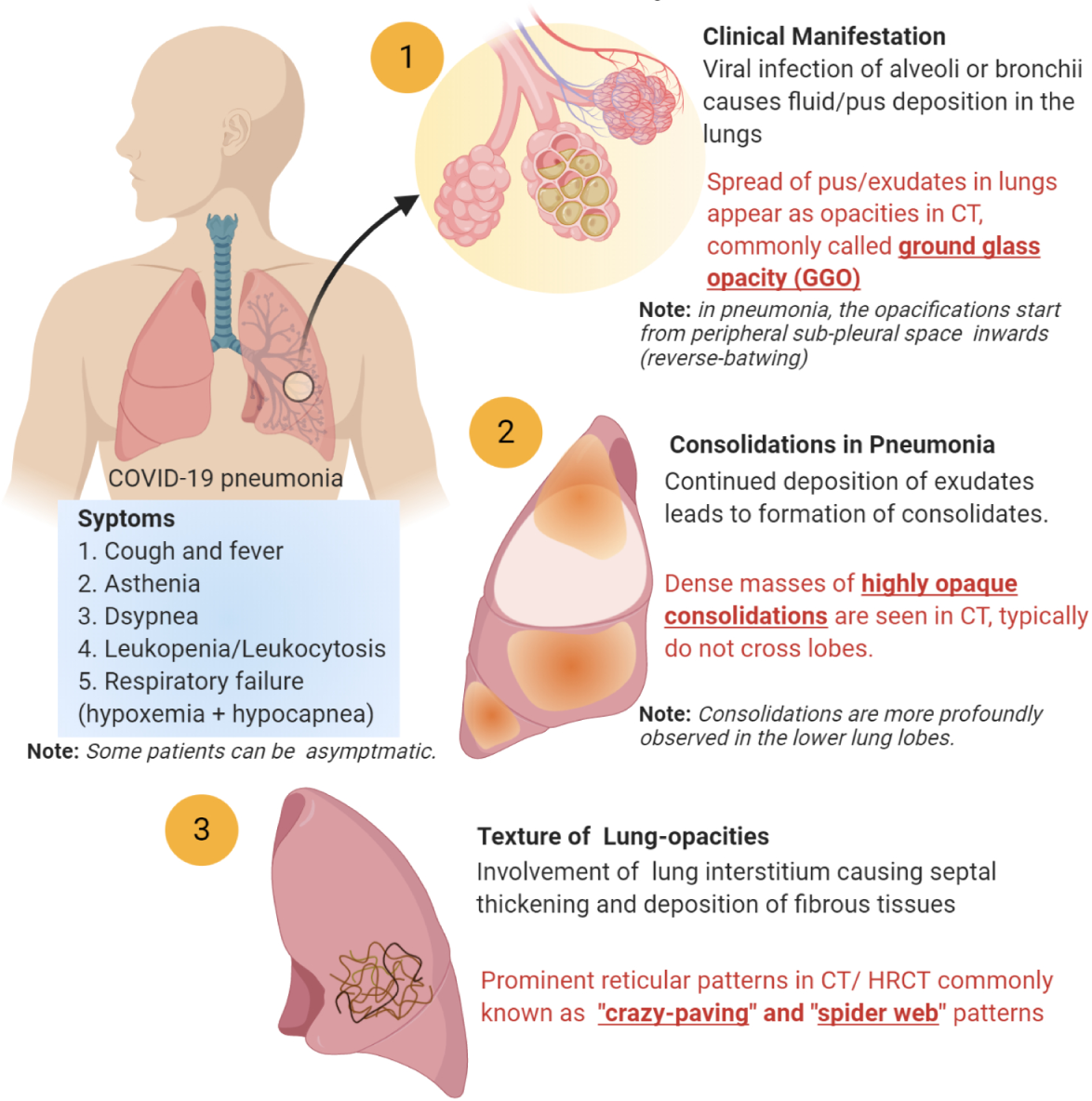
Illustration of pathological features of lung pneumonia in COVID-19 patients along with corresponding CT features.

Due to the entire range of symptoms expressing in the population from asymptomatic to fatal, severity assessment is crucial for effective administration of the right therapeutic drugs as per the patient’s condition [12]. This becomes even more complicated with underlying conditions as some drugs that are otherwise effective for the virus, may have adverse effects on pre-existing conditions. Currently severity assessment is done by symptoms and chemical tests (liver function, pO2, saO2, procalcitonin, troponin, creatinine, blood cell count, inflammatory markers etc.). However most of the specific markers express differently in different stages of the disease [13] and provide the indirect status of the most affected organ, i.e. the lungs.

### Review of Literature

Assessment of 3-class severity (mild, moderate, severe) is crucial to determine the treatment route [14] and is well summarized in the WHO interim guidance on clinical management of COVID-19^2^. Some recent literature have shown chest CT to determine COVID-19 severity qualitatively and quantitatively with good correlation with clinical parameters [15–17]. Further, recent studies have also demonstrated that quantification of lung CT based severity can predict the short-term prognosis of COVID-19 [18, 19]. A number of publications have highlighted methods for determining severity from lung CT images qualitatively, semi-quantitatively, and quantitatively. Schaible et al. proposed two new CT image features —margin sharpness and geographic shape from 108 COVID-19 patients to assess severity [20]. However the effectiveness of these features to stratify severity is yet to ascertained. Yang et al. have analysed 20 segments from chest CT images (with constant CT parameters) manually (by expert) and based on an objective rating of opacification cover in each segment, have assessed mild and severe disease state [21]. Although the evaluation is clinically thorough, the method is expertise-heavy and the requirement for a manual scoring of 20 different segments to obtain a severity score makes prediction of an incoming case cumbersome. Shen et al. on the other hand have used (1) lesion percentage cover and (2) mean lesion density to evaluate 3-class severity in 44 affected individuals [22]. They have used a computer-aided tool to semi-automatically segment lesions and identify the correlation between the two parameters and chest CT pathological features. Although, the computer aided performance correlated well with expert performance, no severity scoring index or predictive performance for classification into severity groups have been shown. Huang et al. have used U-Net deep learning architecture for segmentation of the lung lesions [23], measured the percent opacification cover, and have shown that this feature was statistically different for the four severity classes. The segmentation method has a high performance and is a major highlight in automated assessment of opacification cover. However, the predictive performance of the feature in classifying severity with clear feature thresholds in differentiating the severity groups needs to be explored. Matos et al. quantified the volume of the disease (VoD) as a CT image parameter in addition to 11 other clinical features to determine severity and predict outcome [19] and the VoD quantification, achieved an ROC-AUC (area under the curve) of 0.75. The search for a powerful quantitative feature to assess severity is elusive due to their high variance and heterogeneity as illustrated in a retrospective study involving 4410 COVID-19 patients [24]. A few studies have achieved good accuracy to classify severity in two-classes. Sun et al. using commercial software quantified a number of CT features including the percent lesion cover and the type of deposition to classify 84 patients into severe and non-severe categories with about 91% specificity and sensitivity [15]. On the other hand, Gouda and Yasin quantified CT features of 120 patients including percentage of high and low opacity and total lesion cover using a CT pneumonia analysis algorithm by Siemens Healthineers [16]. With the aid of AI-rad —an artificial intelligence based lesion detection tool they could classify mild and non-mild cases with a sensitivity of up to 90%. The use of AI based lesion detection tools have indeed played a key role for predicting several outcomes like severity and disease progression in COVID-19 [25]. Although a number of published articles have provided severity assessment, very little research is being done to address issues emerging from images sourced from multiple radio-diagnostic centers and hospitals. With the huge clinical burden of COVID-19, it is important to have common assessment features that can cater to images from a variety of imaging units with different CT parameters. Feng et al. performed a multi-center retrospective study on 298 COVID-19 patients wherein they semi-quantitatively estimated severity from CT images using lesion coverage in the lungs. They determined that the CT severity is closely linked with a number of clinical risk parameters and hence discussed how CT based severity measurement can help in risk stratification [17].

A major step in CT image analysis is detection of lung abnormalities. A number of methods ranging from manual to automatic detection are available [26, 27] today. While expert based manual detection incorporates crucial domain knowledge, it is difficult to cater to a huge number of images. Automatic detection methods incorporating advanced image processing and artificial intelligence overcomes this drawback to a large extent but may face problems in delineating borderline cases [28]. Semi-automatic or computer assisted detection methods provide several advantages [29] as they bridge between the two approaches but can still be limiting when the disease burden is very high like that of COVID-19.

### Logical Exposition

Recent literature have correlated chest CT image features with the corresponding pathological disease severity [22, 30]. These gradual emergence of pneumonia attributes —ground-glass opacifications (GGOs), reticular patterns, and dense consolidations [31] in CT images were found to correlate with exudate accumulation and septal thickening/lung fibrosis affecting breathing. Since these pathologies causes a higher absorbance of X-rays (and hence shows a higher CT value) [32], the disease severity should correspond strongly to the lesion gray scale intensity. However, the gray scale-intensity has been found only to be moderately correlated to disease severity [22]. This is because CT scans are often modified among CT centers. Also, individual CT scanners can be set to variably operate at specific CT window settings (level and width) as per user’s requirement. Additionally, the X-ray tube settings, rotation time, pitch, slice thickness etc. are some of the other variables that can affect the perception of lesion ‘density’ in a CT slice and hence severity analyses. Also, image contrast-enhancement for better visualization post-imaging is often performed affecting the CT-values further. Thus a type of normalization of the lesion gray scale intensity is required i.e. independent of the amount of post-processing and parametric variables adopted in CT imaging. Although use of multiple features for severity assessment is important, but multi-variate analysis, use of complex and black-box methods of classification often affect the interpretation of the findings especially for a disease like COVID-19 where images are used to indirectly interpret the pathological severity. Here we have shown an anatomically normalized intensity as a lesion feature and relevant framework to assess severity of lung pneumonia in COVID-19 patients. Comparative evaluation with other methods and features, methodological validations, and relevant thresholds have been also demonstrated.

### Specific Contributions

In this paper, a retrospective study of 102 COVID-19 positive individuals (including multiple follow-ups) has been done to mine the relevant patient history/data to evaluate ground truth severity of the patients. The main contributions of the paper are listed as follows:

i. A CT image feature —*L*_*norm*_ to stratify COVID-19 severity into three classes (mild, moderate, and severe) having linear correlation with disease severity from multi-center image data-set.
ii. A framework for detection of COVID-19 associated pneumonia lesions using manual, computer assisted (semi-automatic), and deep learning-based (fully automatic) methods adapted to quantify *L*_*norm*_. We have not attempted to improve state of the art CT nodule/lesion detection methods but have only adapted the most relevant methods for quantifying the feature *L*_*norm*_.
iii. Quantification of cut-off values with optimum specificity and sensitivity to classify among healthy, mild, moderate, and severe classes.
iv. Quantification of agreement in severity assessment between multiple raters using the feature *L*_*norm*_.
v. Comparison of *L*_*norm*_ with 13 different machine learning based classification methods and against nine other popular image features/feature combinations for both 3-class and 2-class (severe and non-severe) classification of severity.

## 2 Methods

A schematic methodological work-flow is illustrated in Fig. 2.

**Figure 2:**
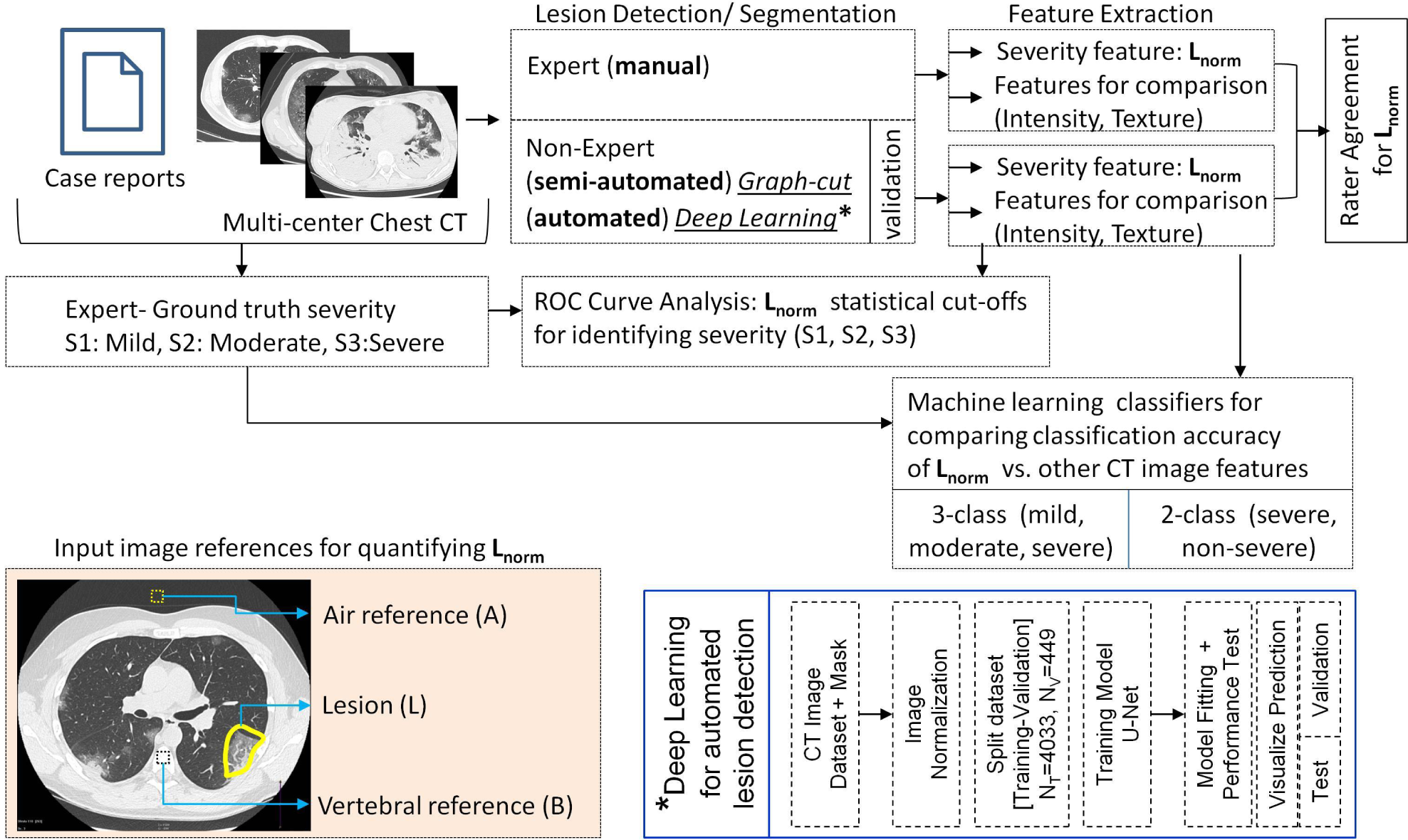
Schematic flow of *L*_*norm*_ feature quantification for severity analysis of COVID-19 affected lungs from multi-center chest CT data. The scheme illustrates the framework for the application and validation of *L*_*norm*_.

### Dataset

All the CT images used for the extraction and validation of *L*_*norm*_ has been taken from the repository of the Italian Society of Medical and Interventional Radiology (SIRM) [33]. A total of 509 CT and HRCT images of 101 COVID-19 positive individuals were taken between the age of 20 to 90 (60% males). All clinical data of the patients were mined and translated to English to be used for determining the ground truth or clinical severity (Supplementary Table S1). No pediatric or COVID-19 negative CT images were included in the study. All clinical annotations given with the cases were used as well. The database includes Multi-center CT scans from 41 different CT/ HRCT imaging centers (Supplementary Table S2).

For automated deep learning based lesion detection, we used four other independent CT image datasets for training the model—(1) Radiopaedia^3^, (2) MedSeg^4^, (3) MosMed^5^ [34], and (4) Coronacases^6^ [35]. All the datasets have corresponding annotated masks to be used for training and validation purposes. All CT images were in ‘.nii’ format, 2D images of size (512, 512) were extracted from them and stored in ‘.npy’ files for training. For the Radiopaedia, Coronacases and MosMed datasets, the images that had blank masks (no visible lesions) were removed from the dataset. The dataset was augmented by adding images which were flipped either horizontally or vertically. Data augmentation was necessary here as deep learning networks perform better with more data, preventing overfitting of the model during training.

### Ground Truth for COVID-19 Severity

All the lung-CT images were evaluated by a radiology expert given the case history, CT images, and corresponding radiological findings in the case-reports to assign three classes based on severity (Supplementary Table S1). The severity was divided into three classes —mild (S-1), moderate (S-2), and severe (S-3) [36] (Figure 3). It is to be noted that S-3 class includes severe and above (including critical) cases.

**Figure 3:**
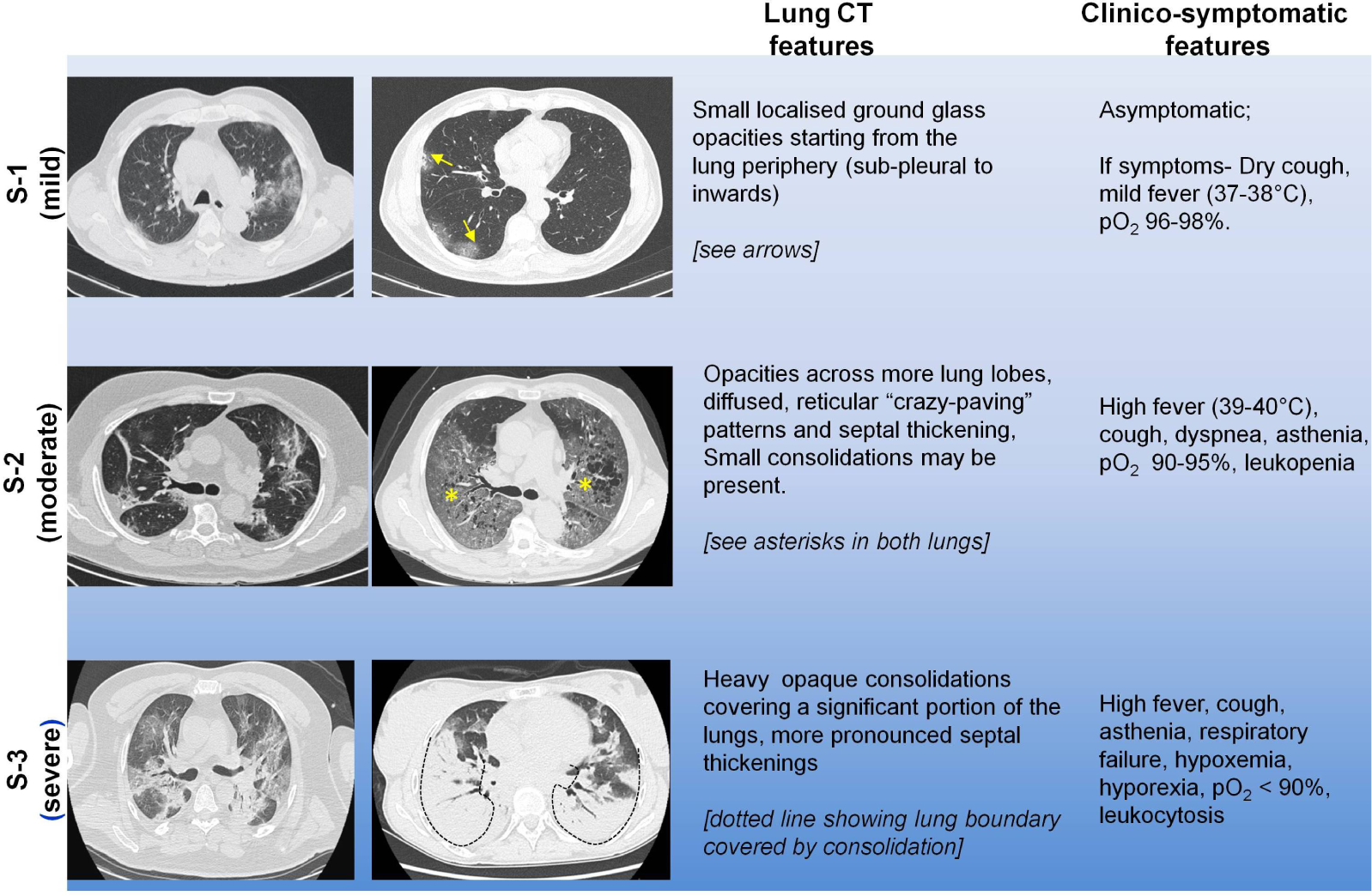
Overview of clinical and chest CT features of pneumonia in COVID-19 patients. Columns 1 and 2 are representative expert assigned chest CT images for the three severity groups. C olumns 3 and 4 are the associated lung-CT features and clinico-symptomatic features. This is a guide to better understand the basis of expert classification (ground truth). *Note:* Features are an approximate guide based on previous literature [22, 30] and does not always correlate completely with severity.

### Lesion Detection

i. Expert (manual lesion area annotation) A radiological expert was provided chest CT images of COVID-19 patients and was asked to manually draw the boundaries of the lung lesions. A total of 180 images were manually annotated by the expert and was used for feature extraction.
ii. Non-expert (semi-automatic lesion detection)
  a. All images were converted to 8-bit gray scale format. No image pre-processing was performed.
  b. Lesion detection-Graph-cut based adaptive region growing algorithm [37] was used to detect the lesions in MATLAB R2019a. Two individuals (without radiological expertise) were briefly trained by an expert to visualize the lesions in the affected lungs. The non-expert individuals provided the initial seed points in the respective foreground (all lesions) and background (rest of the image) and the lesion boundary was detected.
  c. Refinement-The detected lesions were refined by morphological opening to remove the co-detected smaller/thinner bronchial structures and pulmonary vessels in the lung tissue. Some segmentation results using this approach are presented in Supplementary Fig. S1-S3.
iii. Non-expert (automatic lesion detection)
  a. A U-Net model [38] was used to segment the lesions from CT scan images of lungs. It is a neural network architecture specialized to perform better for biomedical image segmentation tasks, where we have limited data and annotations.
  b. Since the datasets used in this work are sourced from a variety of imaging centers, the intensity ranges of the images varied significantly. Thus, histograms were plotted for images of each dataset (Supplementary Fig S4). The values of the pixels were adjusted to lie between the range −1500 to 500. Normalization was done by subtracting the image with the mean pixel value divided by the standard deviation.
  c. The histograms of the lesion masks revealed that the number of pixels corresponding to infections(white pixels) is very little compared to the number of background pixels (black pixels) (Supplementary Fig S4). This is a huge class imbalance and we would need to employ special methods during training to avoid this. All images were resized to a size of (256, 256) for training.
  d. U-Net model: The Python package **segmentation_models**^7^ has been used to create our model as it offers a high-level interface to create and test models quickly. The model has three parts, a backbone, a decoder block and the head. The backbone was essentially a pre-trained model without the last dense layers, which was used as a feature extractor. The **efficientnetb0** model initialized on imagenet weights was used as our backbone. The decoder block consists of 5 upsampling blocks with filter sizes of 256, 128, 64, 32, and 16. The head of our model was a 2D convolutional layer defining the number of output classes (2 in our work) with sigmoid activation.
  e. the training was performed on Google Colab —a free Jupyter notebook environment that runs entirely on the cloud and provides the use of free GPU. The model was trained on 4033 images and validated on 449 images (training-validation: 90-10 split).

### Performance of Lesion Detection/Segmentation

i. For graph-cut method, A binary mask was created from the resultant detected lesions (expert and non-expert) and was used to segment the lesions from the 8-bit gray scale images. The performance of the computer-aided segmentation was validated against the expert labelled lesion area using the Dice similarity coefficient (DSC) or F1 score given by

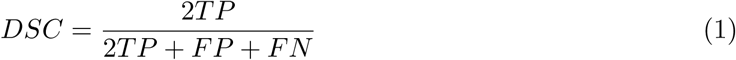

where, TP= True Positive, FP=False Positive. and FN= False Negative. Each connected component was considered as a single lesion. Feature extraction was performed for all the segmented lesions.
ii. For the performance evaluation of the lesion detection by deep learning method, the following steps were taken.
  a. Owing to the class imbalance, we used the focal Tversky loss function [39], which is derived from the dice coefficient (DSC). The Tversky Index (TI) is given by

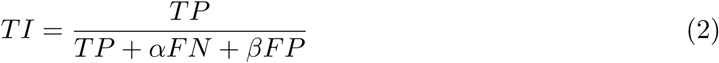

where *α* and *β* are rational numbers. The focal Tversky loss function (FTL) is given by

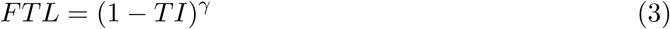

where *γ* is a rational number.
  b. in addition to F1 score (DSC) and Tversky Index, we also used other metrics like precision, recall, and mean IoU (Intersection-Over-Union) given by

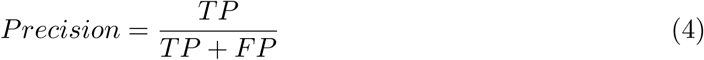

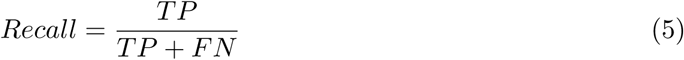

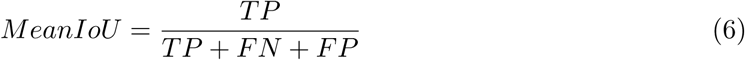
  c. For training we used *α*=0.45, *β*=0.55, and *γ*=0.75. The performance of the lesion detection on the validation set has also been tested with other values of *α, β*, and *γ* listed in Table 1.
  d. The Adam optimizer was used to optimise the loss function as it was found to perform better than SGD, Nadam and Adamax by converging the model faster. It was found that a learning rate of 10^*−*4^ was the best starting point for training the model. The use of higher learning rates gave unstable results, while use of lower learning rates made the model converge slowly. The Keras callback **ReduceLROnPlateau** was used to reduce the learning rate if it did not improve during the last two epochs by a factor of 0.1. A batch size of 8 was used, the model was trained for 40 epochs. The convergence plots are provided in Supplementary Fig S5.

**Table 1:**
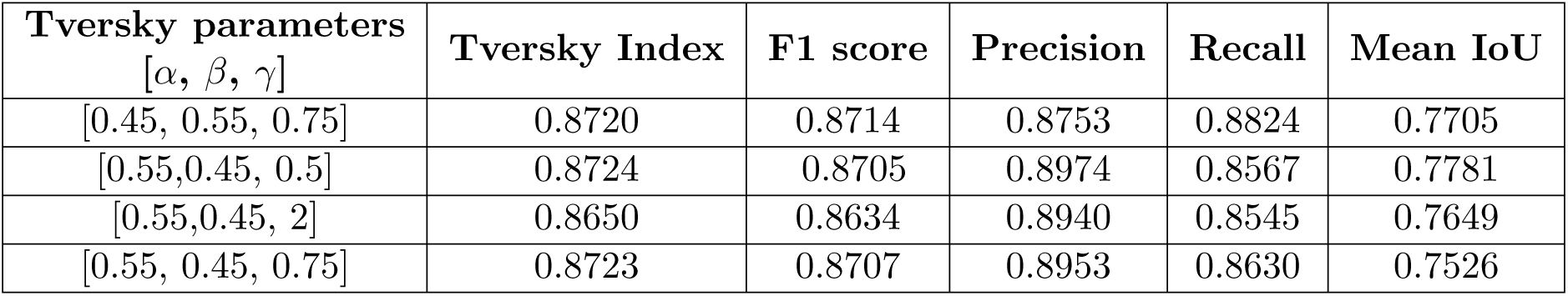
Performance of deep learning based lesion detection predicted on the validation set.

### Feature Extraction and Quantification

Since in CT, the bone has the highest CT value i.e. ∼ 400-1000 Hounsfield units (HU) and the air has the lowest i.e. −1000 HU, the vertebral disk (cancellous region) was taken as the maxima bone reference (*B*) and the air-region exterior to the chest in the same image was taken as the minima air reference (*A*) (Figure 2 colored box). The choice of the vertebral cancellous region was to reduce the CT-value variability within bones. The non-expert after segmenting the lesions gets automatically prompted to position a 30 pixel diameter circle each for the minima (air cavity) and maxima (vertebra) from the same CT image. The program then evaluates the *L*_*norm*_. First, the mean gray intensities of *A, B, L* were calculated from the region using 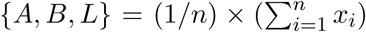, where *x* is the pixel intensity between 0 to 255. Once all the three values (*A, B, L*) are determined, the *L*_*norm*_ was calculated by

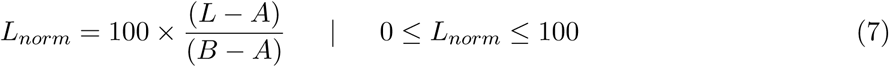

All analyses were performed in MATLAB R2019a. In case of multiple lesions, up to two lesions with the highest *L*_*norm*_ were selected for severity assessment. Based on the values of *L*_*norm*_, the lesion was categorized into three severity classes (S-1, S-2, S-3) using cut-off values evaluated from ROC analyses (discussed in subsequent section).

### Validation

#### i Statistical Analysis

Pearson’s correlation test was employed to evaluate the correlation between *L*_*norm*_ and the mean gray scale intensity of the lesion (L). A total of 163 evaluations from both groups were considered, and the test was performed with two-tailed t-test and 95% confidence interval (CI).

One way ANOVA (with multiple comparisons) was performed to evaluate how well the feature *L*_*norm*_ can delineate the three severity states and the p-value was estimated at 95% confidence interval. A two-tailed t-test was additionally performed to evaluate separation between group pairs.

To identify mild from non-mild cases and severe from non-severe cases, Receiver Operating Characteristic (ROC) curve analysis was performed at 95% confidence interval. The area under the curve as well as the optimum cut-off with the highest combination of sensitivity and specificity was determined. The radiological Lung-CT scores was the ground truth for allocating the individual groups to be delineated.

To evaluate the agreement between and within raters (1 radiology expert and 2 non-experts), *κ*-statistic was used (at 95% CI). All statistical analyses were performed in GraphPad Prism platform.

#### ii Numerical weighted accuracy

The evaluated results of quantified severity assessment using *L*_*norm*_ by an expert and two non-experts was validated against the ground truth disease severity. To obtain the overall weighted percentage accuracy of agreement between the ground truth and *L*_*norm*_ findings, we used:

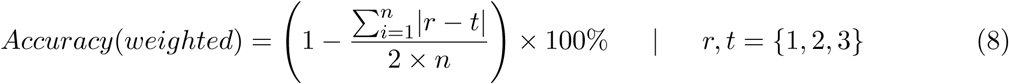

where *n* is the number of cases, *r* = ground truth based severity, *t* =*L*_*norm*_ based severity; 1, 2, 3 correspond to mild, moderate and severe classes.

#### iii Machine Learning for Estimating Severity

To evaluate the computational performance of *L*_*norm*_ to achieve the three-class classification (mild, moderate, and severe), a number of machine learning based classifiers were employed e.g. Decision trees, Naïve Bayes, KNN, Ensemble classifiers etc. *L*_*norm*_ values evaluated from multiple lesions by one expert and two non-experts which constituted a total of N=248 for classification. For classification, the sample size was partitioned randomly in a ratio of 60:40 (training:testing). A number of intensity (gray-scale intensity, lesion standard deviation) and Gray level co-occurence matrix (GLCM) texture features (angular moment, contrast, correlation, inverse difference, and entropy) were measured for the same set of lesions to compare their classification performance with *L*_*norm*_. The classifiers were trained in MATLAB R2019a and were 10-fold cross validated in order to avoid over fitting by the classification models. Principle component analysis (PCA) was used for dimensional reduction in multivariate trained models with intensity and texture features. The testing accuracy of the best classification model was then determined.

## 3 Results

The CT image dataset presented key images, patient history, clinical, and radiological findings. After processing the entire dataset and filtering cases as per the inclusion/exclusion criteria, we determined that the images used in this study were procured from 41 different imaging centers/hospitals spread across Italy (Supplementary Table S2). The severity index —S-1, S-2, and S-3 were assigned to the severity conditions—mild, moderate, and severe (Fig. 3, Supplementary Table S1). We found that for a multi-center CT data with high variation between images,the *L*_*norm*_ value has almost no correlation with the primary variable it is derived from i.e. the mean lesion intensity (Figure. 4). Furthermore, upon comparing the *L*_*norm*_ of healthy lung tissue and COViD-19 lesion, it was observed that the feature could distinguish all COVID-19 lesions including even the mild lesions from healthy lung with 100% sensitivity and specificity (Figure. 4 b-e).

**Figure 4:**
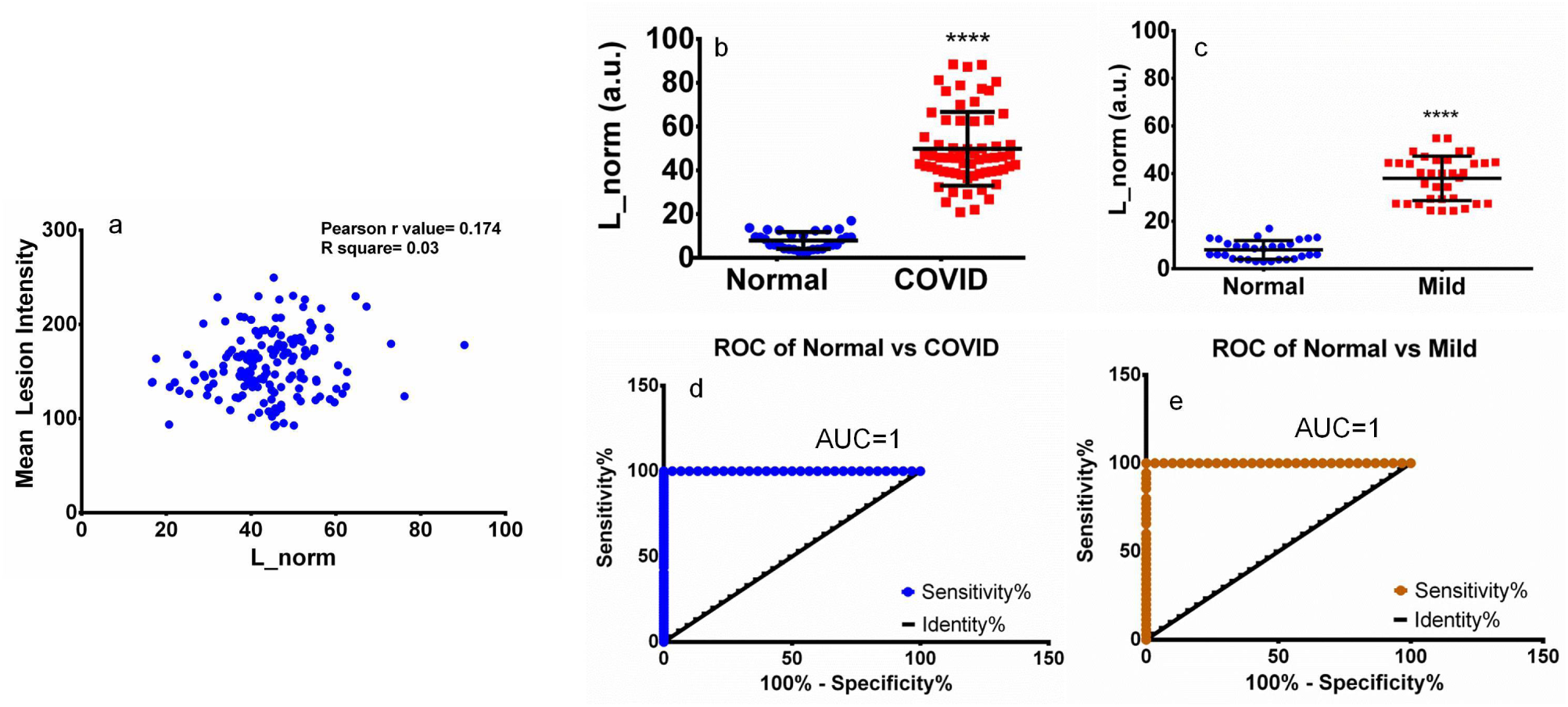
*L*_*norm*_ to delineate healthy from pneumonia affected lungs. (a) Pearson’s correlation test showing that the mean lesion intensity has almost no correlation with *L*_*norm*_ (N=163). (b) plot shows that *L*_*norm*_ of healthy and COVID-19 lesions are significantly distinct (N=94, p-value<0.0001, two-tailed t-test), (c) plot shows that *L*_*norm*_ of healthy and mild COVID-19 lesions are also significantly distinct (N=65, p-value<0.0001, two-tailed t-test), (d and e) are the ROC curve of normal vs COVID-19 and normal vs mild COVID-19 showing *L*_*norm*_ can delineate healthy and COVID affected lung lesions with 100% specificity and sensitivity.

### Lesion Detection and Feature Extraction

The lesions were detected using graph-cut region growing initiated by user-defined seed points for foreground and background followed by morphological opening (Fig. 5). We observed that the segmentation method detected lesions that are —small and faint (Fig. 5a-c), multiple with size variations (Fig. 5e-g), periphery localized (Fig. 5i-k) as well as pan-lung (Fig. 5m-o) with significant overlap with expert-determined lesion area (Fig. 5d,h,l,p). A few more results to illustrate the segmentation performance of CT images from multiple centers are shown individually for mild, moderate and severe cases in Supplementary Fig. S1, S2, S3. A Dice similarity coefficient (DSC) of 0.887(*N* = 180) was found when the segmentation results were compared against expert-marked lesion area.

**Figure 5:**
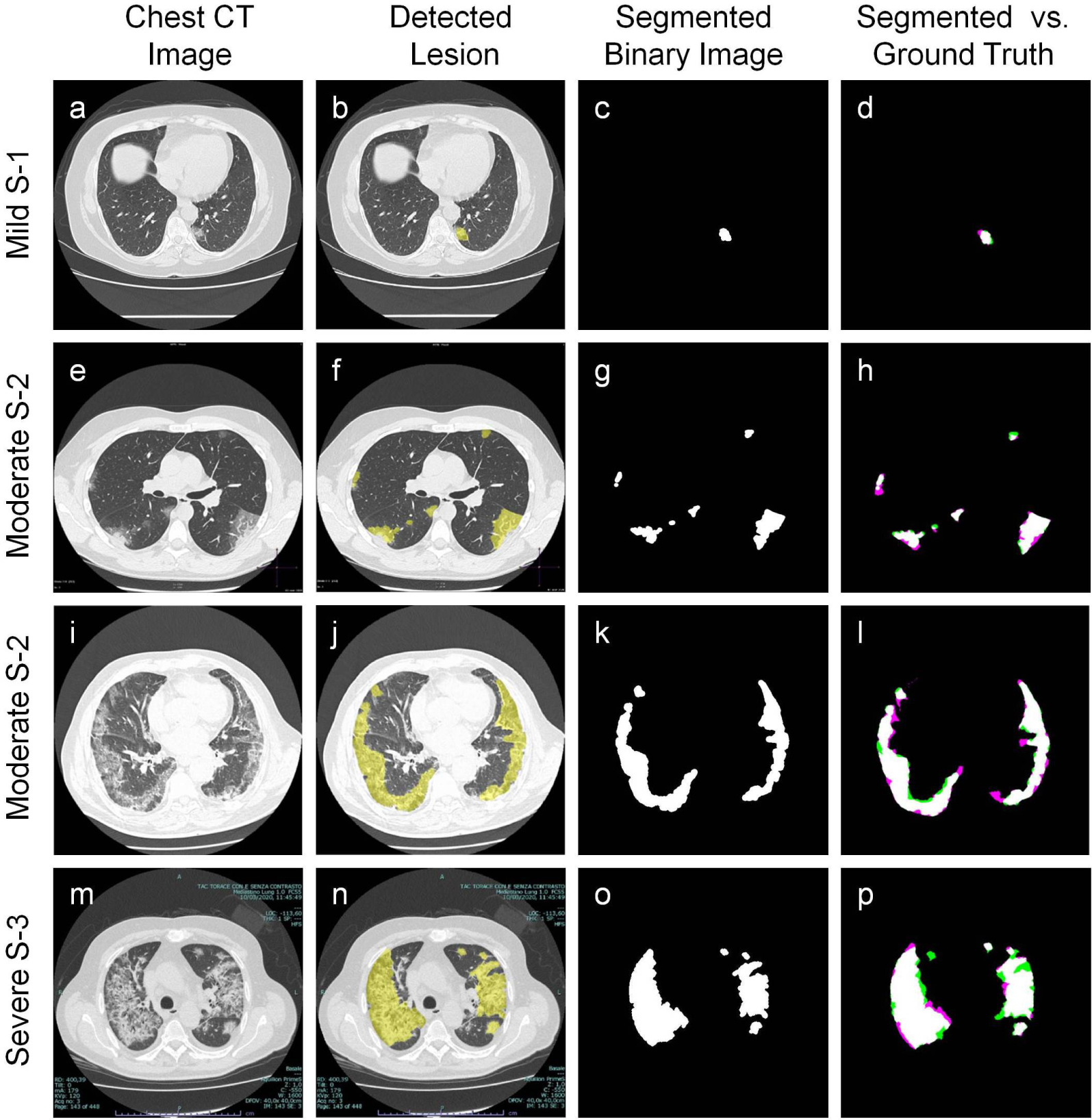
Semi-automated lesion detection of pneumonia-affected lungs in COVID-19 patients. (a,e,i,m) original chest CT images with lesions of different size, number, and localization, (b,f,j,n) lesions detected by the graph-cut based lesion detection, (c,g,k,o) binary images of the segmented lesions, (d,h,l,p) overlay of segmented result and expert-marked lesion area. The white area depict fully overlapped region, the orange and magenta areas depict exclusively segmented and expert-labeled regions respectively.

The deep-learning based method also effectively detected the lesions (Figure 6) with significantly high accuracy i.e. DSC of 0.87 (N=449). The different performance metric scores for lesion detection using deep learning are listed in Table 1. The convergence plots for evaluating performance are available in Supplementary Fig. S5. Detection of different types of lesions are also shown in Supplementary Fig. S6.

**Figure 6:**
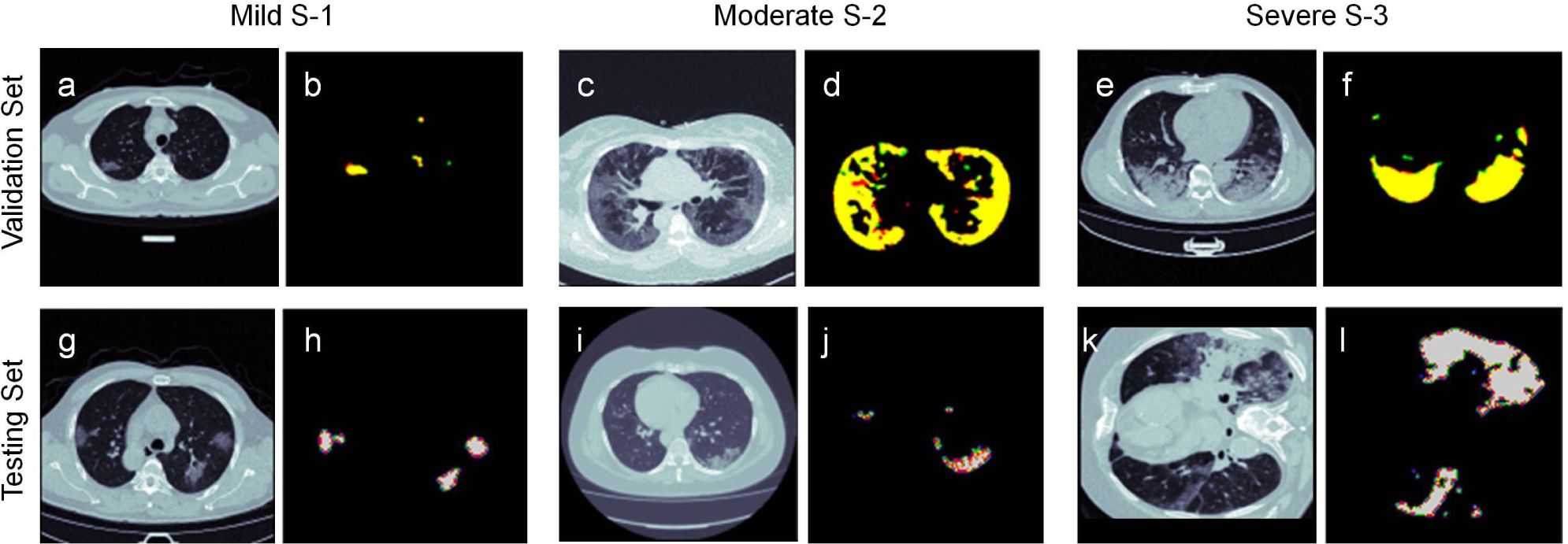
Automated detection of lesions of pneumonia-affected lungs in COVID-19 patients. (a,c,b) are CT images from the validation set, representing mild, moderate and severe pathology, (b,d,f) are the corresponding predicted lesions. The color code is—Yellow=true positive, Red=false-positive, Green=false-negative. (g,i,k) are CT images from the testing set, representing mild, moderate and severe pathology, (h,j,l) are the corresponding predicted lesions.

Thus, it was noted that both the non-expert based methods have similar performance in detecting the lesions.

### Determination of Thresholds to Delineate the Severity Conditions

*L*_*norm*_ feature values to identify the severity was determined using Equation 7 from segmented lesions shown in Fig. 2. The ground truth score determined by the in-house experts after going through the database reports (see general guidelines in Fig. 3) along with the evaluated *L*_*norm*_ values are given in Supplementary Table S1. The distribution of *L*_*norm*_ values in each of the ground truth categorized severity groups from the heuristic inputs of both experts (Fig. 7a) and detected inputs of non-experts (Fig. 7d) shows the features are statistically distinct for the three severity stages (p-value < 0.0001, 95% CI). Since the detection performance of both non-expert semi-automatic (graph-cut) and non-expert automatic (deep learning) lesion detection was similar, their *L*_*norm*_ evaluations were combined under non-expert category.

**Figure 7:**
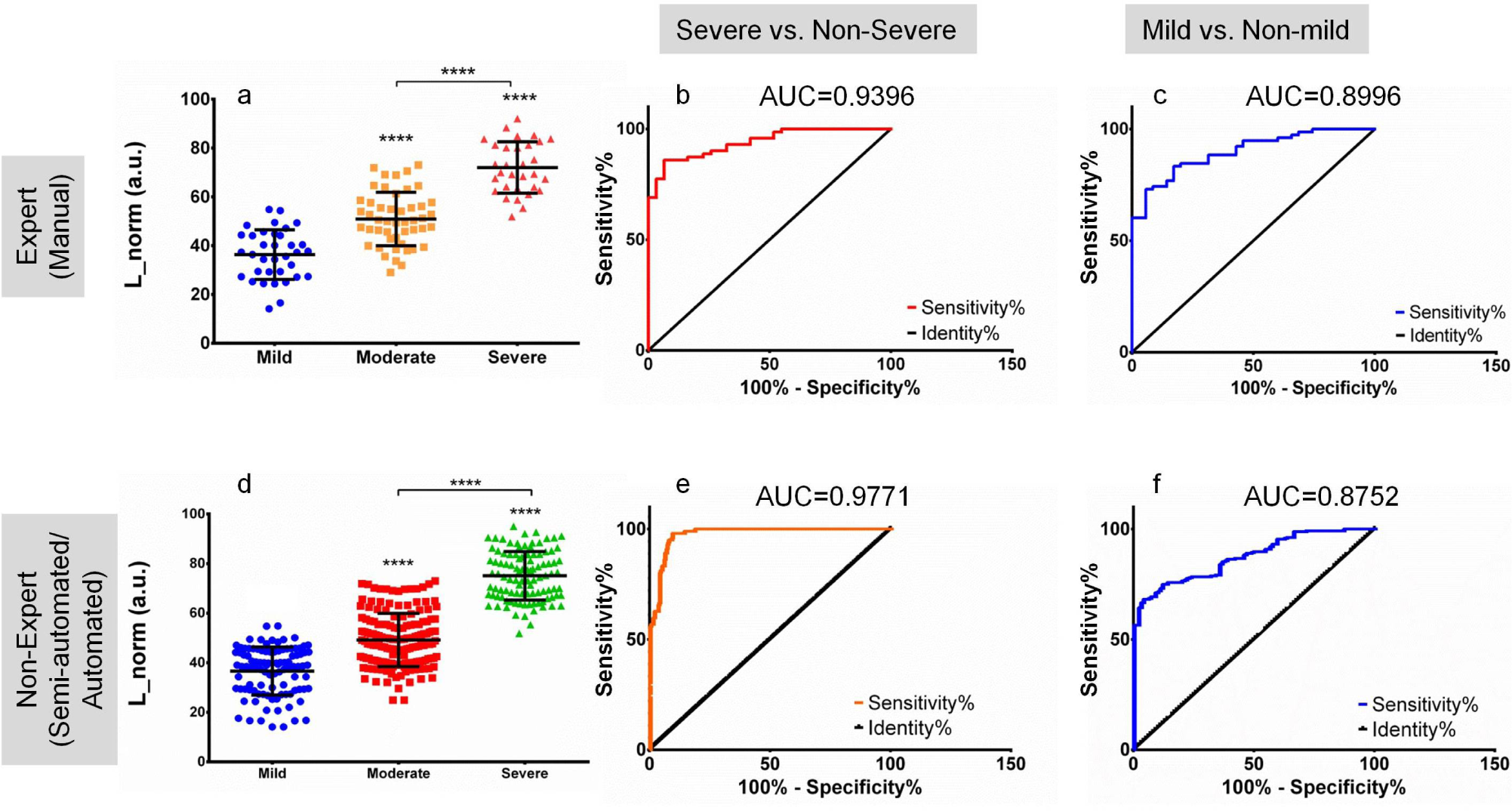
Statistical analyses for determining *L*_*norm*_’s potential in severity assessment. (a-c) are statistical findings from one expert annotations (N=191) while (d-f) are from non-expert annotations (N=425). (a,d) shows *L*_*norm*_ is well separated for the three severity cases,between all groups p-value <0.0001, (b,e) ROC analyses for classifying severe from non-severe class, (c,f) ROC analyses for classifying mild from non-mild class. Statistical analyses in (a) and (d) is done with one-way ANOVA, followed by Tukey’s post-hoc test for multi-group comparison.

The optimum cut-off values of *L*_*norm*_ to identify the three stages were determined by the ROC analysis (Fig. 7b,c,e,f). Based on the best sensitivity and specificity combinations from the ROC analysis (from both expert and non-expert data) to delineate severe from non-severe (sensitivity/specificity-92.26%/95.1%) and mild from non-mild cases (sensitivity/specificity - 75%/87.3%), we determined the cut-off for each of the three categories and normal lungs (Table. 2). Both expert and non-expert assigned demarcations had a high accuracy for delineating severe from non-severe cases with ROC area under the curve (AUC) of 0.939 and 0.977 (Fig. 7 b,e). Similarly, results from expert annotations (AUC=0.899) were comparable to non-expert (AUC=0.875) for delineating mild from non-mild cases (Fig. 7 c,f). Thus, in conclusion, the feature *L*_*norm*_ is found to be well-differentiated for the three severity conditions in a multi-center data across experts and non-experts.

### Agreement Among Human Raters for Severity Assessment

To determine the accuracy of chest CT feature *L*_*norm*_ based severity to match the annotated ground truth we evaluated the weighted accuracy as per equation 8. The accuracy was measured using the expert derived *L*_*norm*_ to assess the closeness of the radiological (expert) measure to a multi-factor ground truth (symptoms, clinical findings, radiological findings etc.). The severity classes were assigned based on the thresholds determined in the previous section (Table 2). The weighted accuracy for the 3-class severity classification was determined to be 86.88%.

**Table 2:** Normalized lesion intensity *L*_*norm*_ for each severity class.

| Severity Score | $L_{norm}$ |
| --- | --- |
| Normal Lung | 0 - 18.86 |
| S-1 (mild) | 18.87 - 46.06 |
| S-2 (moderate) | 46.07 - 61.84 |
| S-3 (severe) | 61.85 - 100 |

To further determine the agreement between raters and dependency of expertise levels and segmentation method, the statistical *κ*-scoring was done involving two non-experts and one expert (Table 3). Both the non-expert intra-rater agreements were the highest (*κ* score 0.965) showing the fidelity of the non-expert lesion detection methods used. The agreement between expert and non-expert ranged from substantial to almost perfect [40]. In conclusion, the high agreement between and within raters demonstrates the reliability and reproducibility of the feature *L*_*norm*_.

**Table 3:**
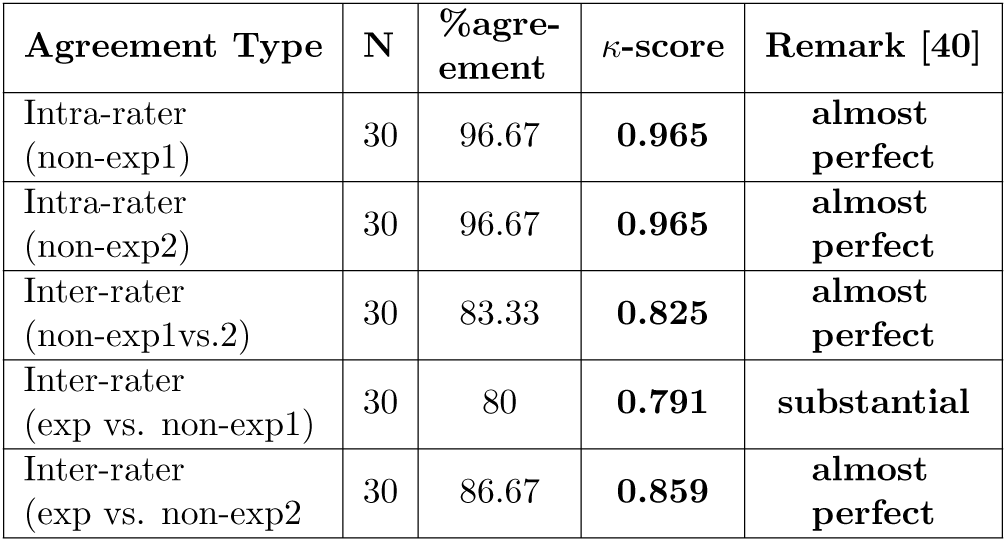
*κ*-statistic to evaluate the agreement of expert and non-expert raters to assign one of the 3 severity classes (mild, moderate, and severe) based on ROC-derived optimum *L*_*norm*_ threshold.

### Machine Learning for evaluating classification accuracy

To establish that *L*_*norm*_ can effectively classify severity in three classes without exclusively assigning optimized cut-offs (like from ROC analysis), we employed machine learning classifiers. A number of classification models were used for training with *L*_*norm*_ values (Table 4). Additionally, we used different standard features of image gray-level intensity and texture associated with the chest CT lesion images to compare their classification performance with *L*_*norm*_. Severity classes (S-1, S-2, and S-3) were assigned as per the annotated ground truth associated with the individual cases for training the models. It was found that the feature *L*_*norm*_ had the highest classification accuracy among all the other individual as well as compound (multi-variate) features for all the classification models (88.2%). Among the classification models, the Decision Tree was determined to have the highest classification accuracy, followed by KNN and ensemble-learning models (Boosted and bagged trees). In conclusion, *L*_*norm*_ alone has a superior 3-class classification performance as compared to the standard intensity and texture features.

**Table 4:**
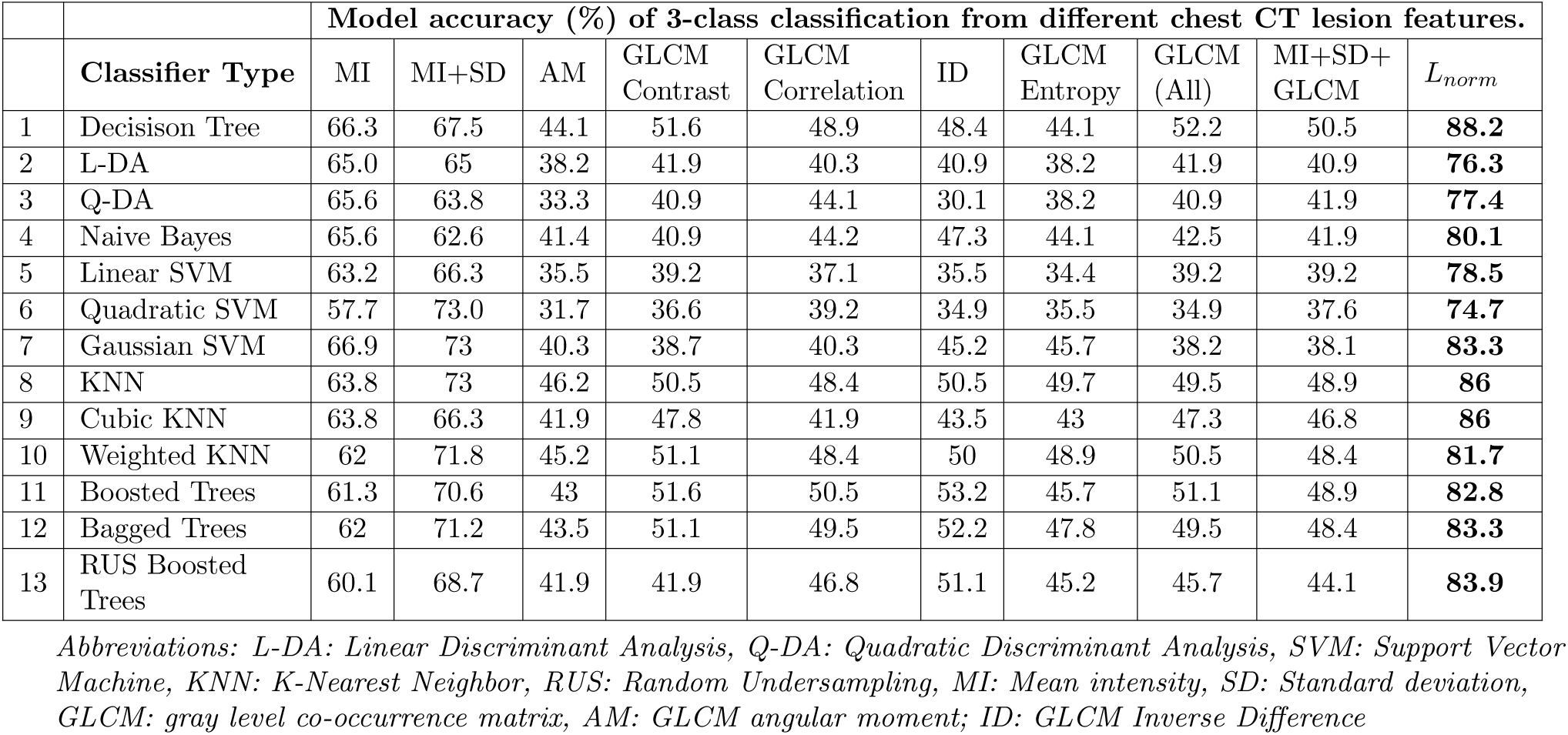
Comparison of classification accuracy based on various popular classification models to achieve the 3-class classification using different chest CT image features vs. *L*_*norm*_

|  |  | Model accuracy (%) of 3-class classification from different chest CT lesion features. |  |  |  |  |  |  |  |  |  |
| --- | --- | --- | --- | --- | --- | --- | --- | --- | --- | --- | --- |
| | Classifier Type | MI | MI+SD | AM | GLCM Contrast | GLCM Correlation | ID | GLCM Entropy | GLCM (All) | MI+SD+GLCM | $L_{norm}$ |
| 1 | Decisison Tree | 66.3 | 67.5 | 44.1 | 51.6 | 48.9 | 48.4 | 44.1 | 52.2 | 50.5 | **88.2** |
| 2 | L-DA | 65.0 | 65 | 38.2 | 41.9 | 40.3 | 40.9 | 38.2 | 41.9 | 40.9 | **76.3** |
| 3 | Q-DA | 65.6 | 63.8 | 33.3 | 40.9 | 44.1 | 30.1 | 38.2 | 40.9 | 41.9 | **77.4** |
| 4 | Naive Bayes | 65.6 | 62.6 | 41.4 | 40.9 | 44.2 | 47.3 | 44.1 | 42.5 | 41.9 | **80.1** |
| 5 | Linear SVM | 63.2 | 66.3 | 35.5 | 39.2 | 37.1 | 35.5 | 34.4 | 39.2 | 39.2 | **78.5** |
| 6 | Quadratic SVM | 57.7 | 73.0 | 31.7 | 36.6 | 39.2 | 34.9 | 35.5 | 34.9 | 37.6 | **74.7** |
| 7 | Gaussian SVM | 66.9 | 73 | 40.3 | 38.7 | 40.3 | 45.2 | 45.7 | 38.2 | 38.1 | **83.3** |
| 8 | KNN | 63.8 | 73 | 46.2 | 50.5 | 48.4 | 50.5 | 49.7 | 49.5 | 48.9 | **86** |
| 9 | Cubic KNN | 63.8 | 66.3 | 41.9 | 47.8 | 41.9 | 43.5 | 43 | 47.3 | 46.8 | **86** |
| 10 | Weighted KNN | 62 | 71.8 | 45.2 | 51.1 | 48.4 | 50 | 48.9 | 50.5 | 48.4 | **81.7** |
| 11 | Boosted Trees | 61.3 | 70.6 | 43 | 51.6 | 50.5 | 53.2 | 45.7 | 51.1 | 48.9 | **82.8** |
| 12 | Bagged Trees | 62 | 71.2 | 43.5 | 51.1 | 49.5 | 52.2 | 47.8 | 49.5 | 48.4 | **83.3** |
| 13 | RUS Boosted Trees | 60.1 | 68.7 | 41.9 | 41.9 | 46.8 | 51.1 | 45.2 | 45.7 | 44.1 | **83.9** |

Abbreviations: L-DA: Linear Discriminant Analysis, Q-DA: Quadratic Discriminant Analysis, SVM: Support Vector Machine, KNN: K-Nearest Neighbor, RUS: Random Undersampling, MI: Mean intensity, SD: Standard deviation, GLCM: gray level co-occurrence matrix, AM: GLCM angular moment; ID: GLCM Inverse Difference

### 3-class vs. 2-class classification

In many scenarios delineation of severe to critical cases are more relevant and therefore we additionally showed the performance of *L*_*norm*_ for classifying severe from the non-severe class. Upon investigating the results of 3-class classification models (Fig.8 a-c) it was seen that most of the errors in the model were concentrated in the mild and moderate classification (Fig.8 a,c). The confusion matrix of the trained Decision Tree model (Fig.8 c) revealed that none of the severe cases were mis-classified as mild or moderate. Thus we re-trained a decision tree model with only two classes i.e. severe and non-severe(Fig.8 d-f). Both mild and moderate cases (S-1 and S-2) were considered as non-severe here. This two-class classification showed a much higher AUC (0.99) and accuracy as compared to the 3-class classification (training (N=148) 98.9%, testing (N=100) 100%). Additionally, the summary of the model accuracy for the 2-class classification for different trained models is summarized in Table 5 highlighting the enhanced performance compared to the 3-class classification.

**Table 5:**
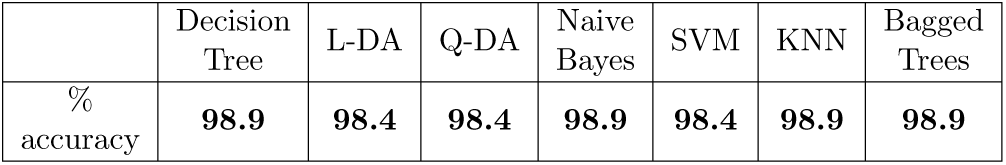
Accuracy of classification models to achieve the 2-class classification (severe and non-severe) using *L*_*norm*_.

## 4 Discussions

The emergence of CT imaging of lungs as an important tool for identifying severity, motivated the quest of a single feature that can be used across multiple imaging centers to perform severity classifications. The images in the dataset were captured from 41 different radiology units and expressed high variations in image quality and contrast (Supplementary Fig. S1, S2, and S3). A number of imaging parameters (discussed previously) determine the intensity of the visualized lesion. Further, contrast adjustment during imaging is a common practice among radiologists to make distinct identification of affected lung regions, and in many practical situations, raw images may not be available or retrievable. To this aim, the feature *L*_*norm*_ does not require raw or pre-processing of the enhanced images. This is essentially due to the inherent anatomical normalization with normalizing elements depending only on the same image-slice as the lesion. This ensures the reproducibility of the method across a range of CT units.

In the recent publications it was shown that mean lesion intensity does not have a very high correlation with the severity features of the lungs [22]. However, although *L*_*norm*_ is primarily derived from the same lesion intensity it has a superior performance in classifying severity. This is because *L*_*norm*_ normalizes large variations among images and is independent of the post-processing and imaging parameters. The amount of this variation is clear with almost negligible correlation between mean lesion intensity and *L*_*norm*_ (Figure 4).

The increased clinical burden of COVID-19 has expedited the necessity of assessing severity of individuals affected with the virus and allocation of radiological expert can be challenging. With a high agreement in severity assessment between the experts and non-experts, the method can be implemented by non-expert staffs with little training for routine evaluation of severity. The feature *L*_*norm*_ shows high performance in delineating severe cases from non-severe ones by both expert and non-expert annotations(Fig. 7). Additionally *L*_*norm*_ showed a similar performance in stratifying the different classes between expert and non-expert based quantification (Fig. 7). Therefore, it can be concluded that the expert dependency is relaxed substantially.

The chest CT gray scale intensity clinically translates to the increase in pathological deposition of exudates along with tissue involvement [31]. As *L*_*norm*_ captures the chest CT intensity, its linear increase, correlates with the increase in disease severity. Besides, the linearity of *L*_*norm*_ reduces the co-dependence of other image features and multi-variate classifier training in order to achieve better classification results. Although specific cut-offs have been provided for classification, it needs to be noted that severity is more continuous than a discrete class bounded by a cut-off margin. Thus, the linear dependence of the *L*_*norm*_ values with the disease severity makes interpretation of the disease condition easier and non-discrete. This is also the reason why even for weak classification learners like Decision Tree, the performance of the *L*_*norm*_ is the highest. This reduces the computational complexity as well.

The different classification models that were employed to classify severity showed that the severity misclassifications are rarely seen when identifying severe cases in the 3 class-classification. Furthermore, no cases were misclassified more than one level of severity i.e. mild cases can be misclassified as moderate but never as severe. This not only shows the fidelity of the feature but ensures almost no severe cases to be misclassified. This became apparent when the classification models were trained for a 2-class severe vs non-severe classification (Fig. 8). The 2-class classification not only showed an extremely high AUC (0.99) but a very high classification accuracy across all the classifiers (Table 5).

**Figure 8:**
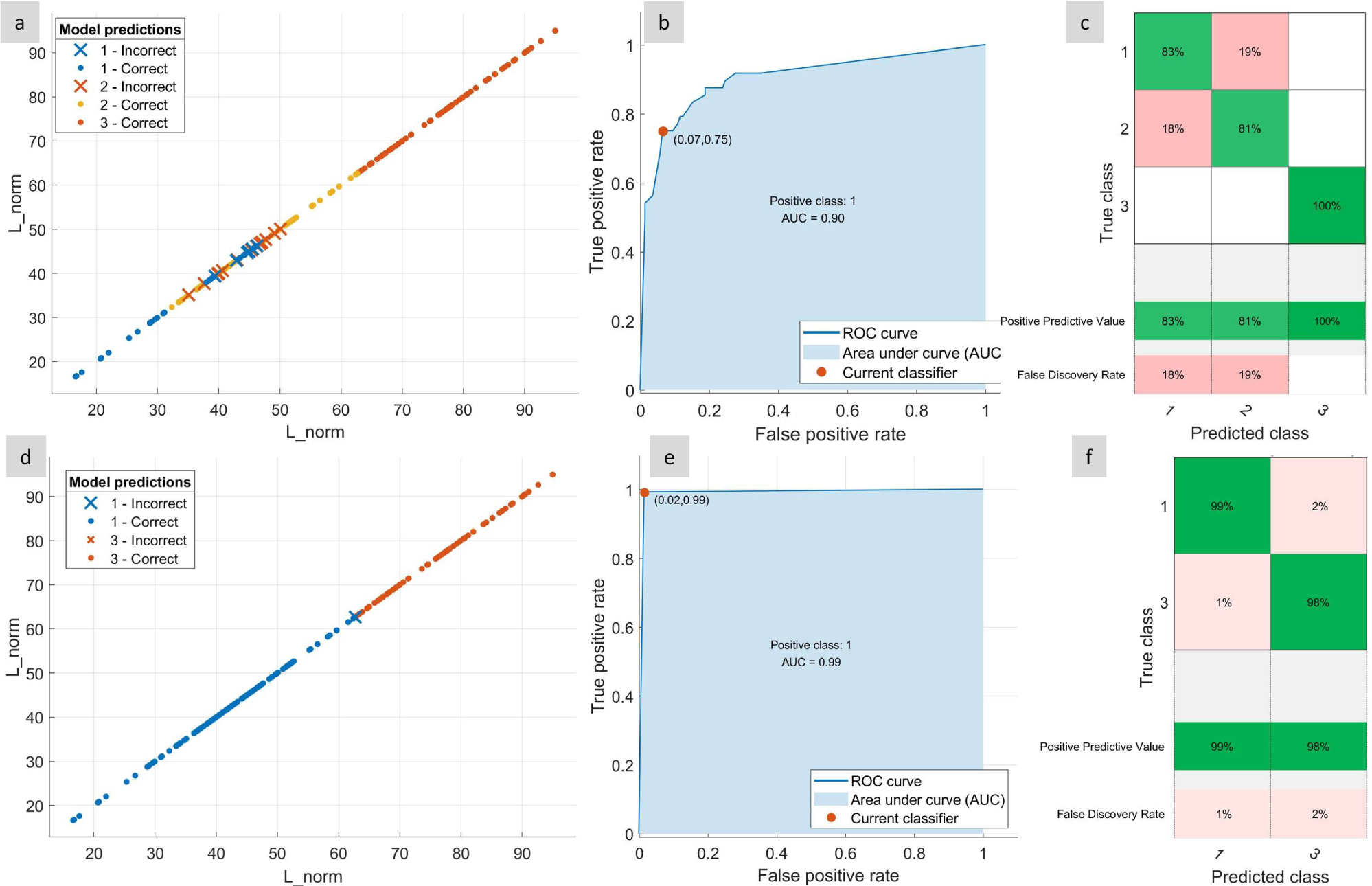
Evaluation of *L*_*norm*_ for 3-class and a 2-class (severe and non-severe) classification using a Decision Tree model. (a) Classification performance for a 3-class 1. mild, 2. moderate, and 3. severe classification (training N=150), (b) corresponding ROC curve and (c) confusion matrix showing no mis-classification for severe cases, (d) performance of 2-class classification 1. non-severe and 2. severe, (e) corresponding ROC curve and (f) confusion matrix showing the positive predictive value and false discovery rate.

The demonstrated quantitative severity assessment from chest CT in COVID-19 positive individuals, if implemented properly can help in managing the patients and provide the necessary treatment to reduce mortality and side-effects. Here we have outlined a scheme of general clinical work flow to demonstrate how the patient management can be done to incorporate the method (Figure 9).

**Figure 9:**
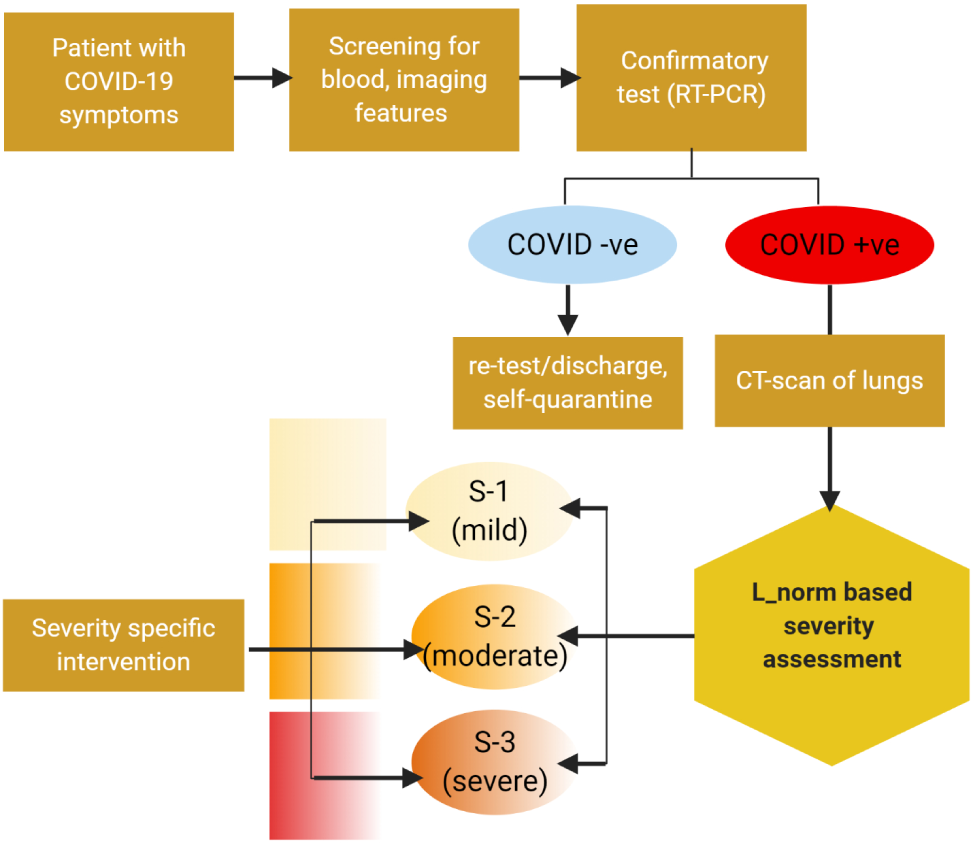
A proposed workflow of resource utilization and incorporation of severity assessment in diagnosis to therapy pipeline of COVID-19 disease in clinical settings.

## 5 Conclusion

To summarize, the article illustrates the lung-CT feature of COVID-19 patients to evaluate their severity quantitatively and group them in three classes of severity —S-1(mild), S-2(moderate), and S-3(severe) using the feature *L*_*norm*_. This feature helps identification of severity groups which can help in therapeutic decision making for reducing risks and mortality in such a wide pandemic. The simplicity of the method along with high agreement score makes it a potential tool to be incorporated in the clinical diagnosis-therapy pipeline for management of COVID-19.

## Data Availability

All data and codes can be made available upon request.

## Footnotes

1 https://www.sirm.org/category/senza-categoria/covid-19/

2 WHO reference number: WHO/2019-nCoV/clinical/2020.5

3 https://radiopaedia.org/playlists/25887

4 http://medicalsegmentation.com/covid19/

5 https://mosmed.ai/

6 https://www.kaggle.com/andrewmvd/covid19-ct-scans

7 https://github.com/qubvel/segmentation_models

