## Supplementary Table, Supplementary Fig for "A Quantitative Lung Computed Tomography Image Feature for Multi-Center Severity Assessment of COVID-19"

### SUPPLEMENTARY INFORMATION

### SUPPLEMENTARY FIGURES

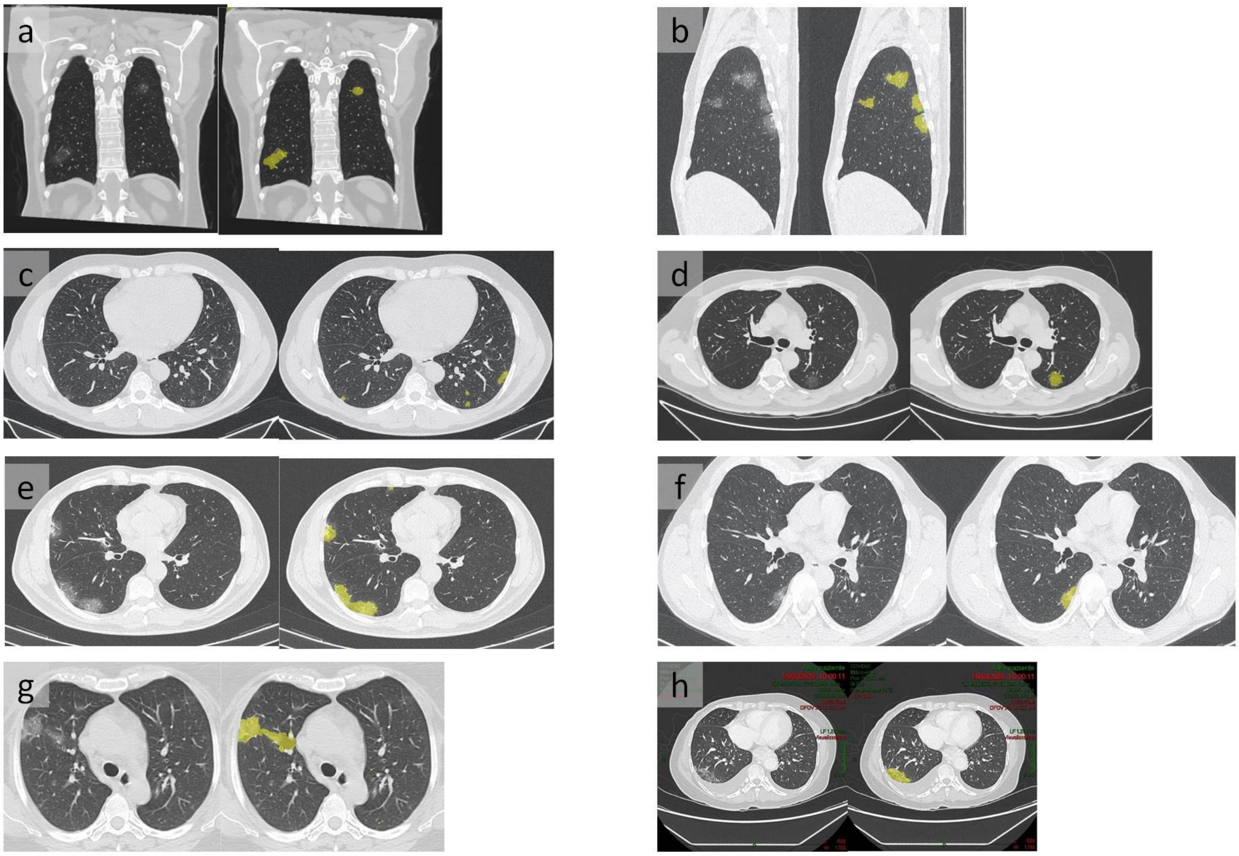


Figure S1: Illustration of multi-center lung CT lesion detection for mild (severity S-1) COVID-19 associated pneumonia..

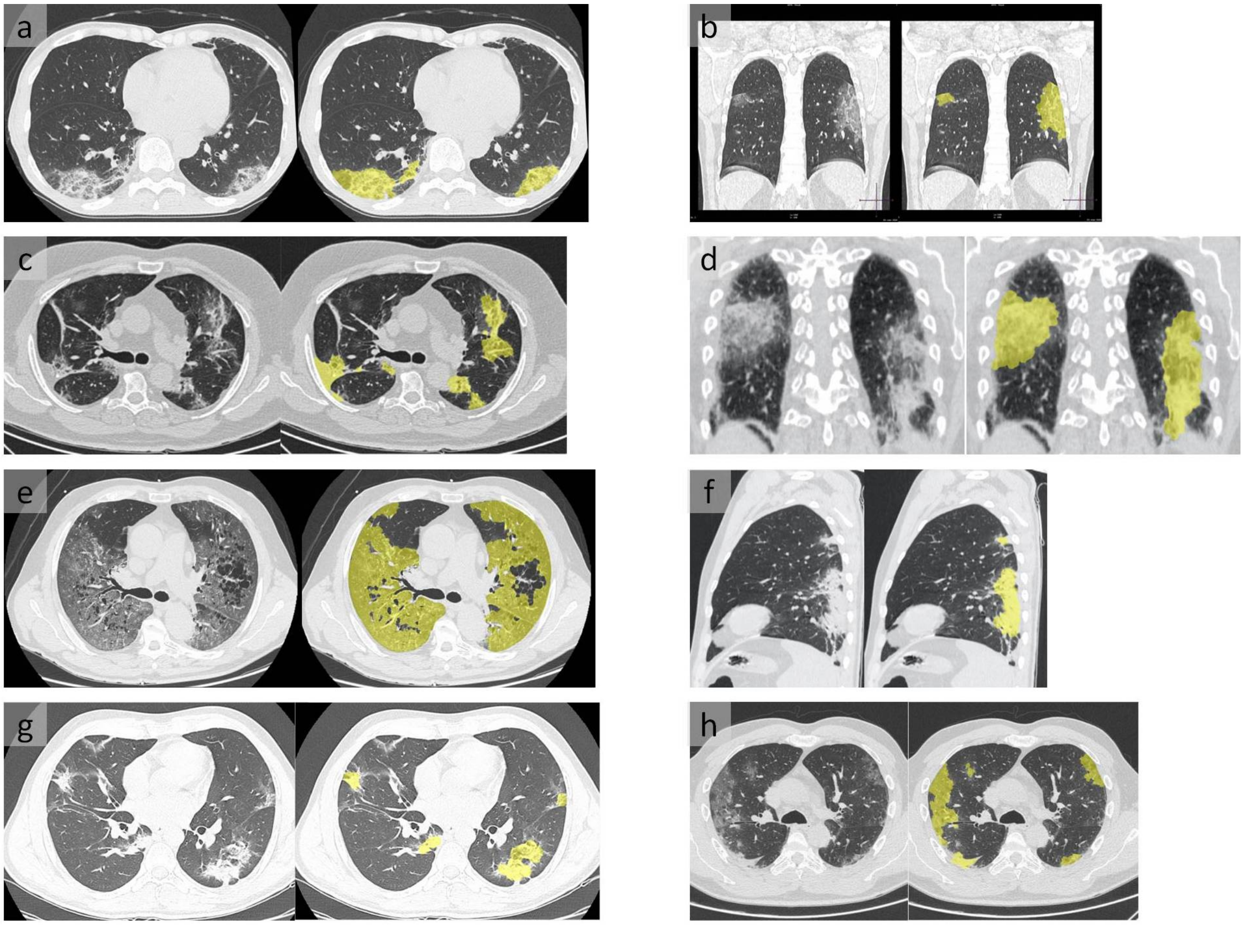


Figure S2: Illustration of multi-center lung CT lesion detection for moderate (severity S-2) COVID-19 associated pneumonia.

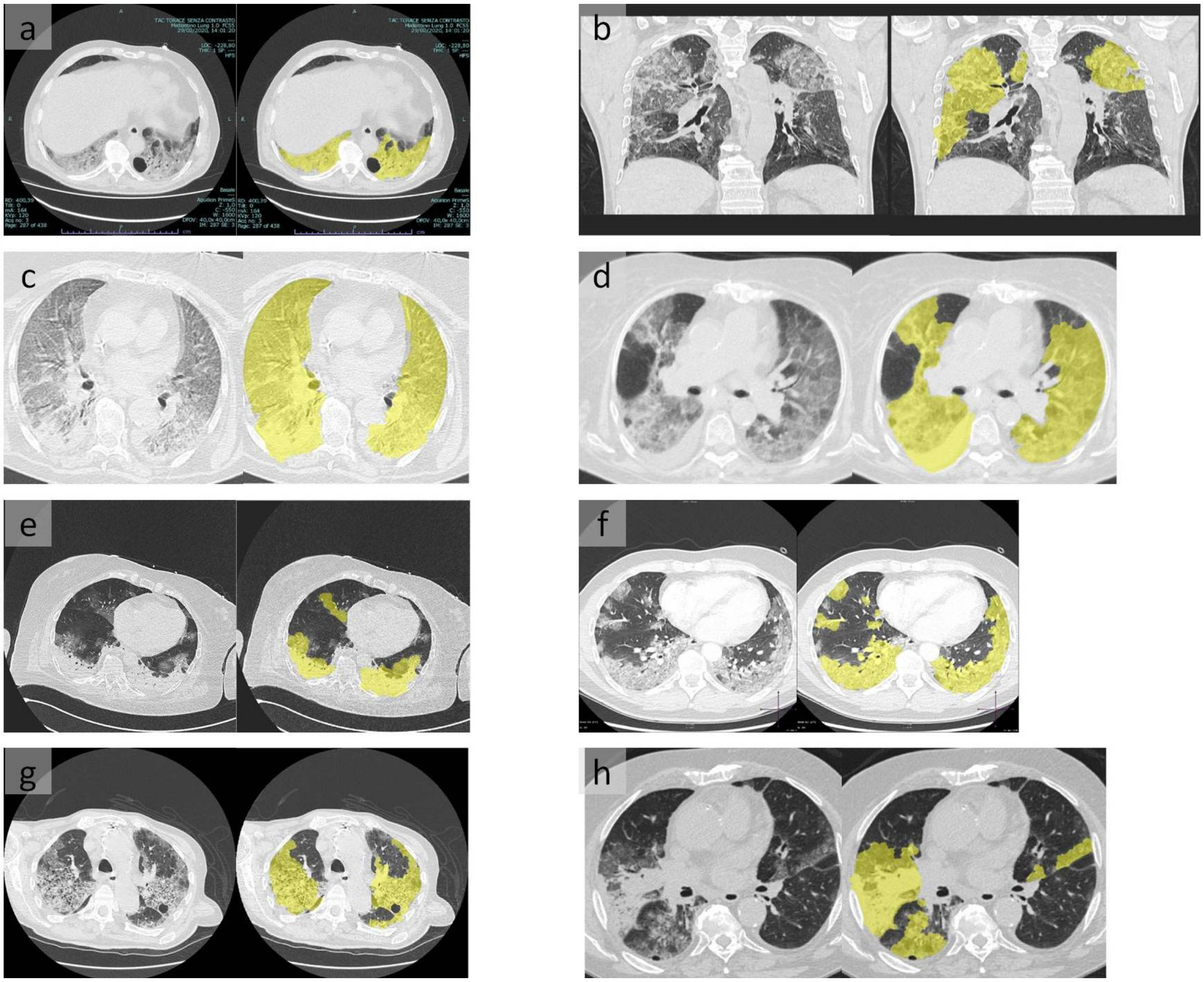

Figure S3: Illustration of multi-center lung CT lesion detection for severe (severity S-3) COVID-19 associated pneumonia.

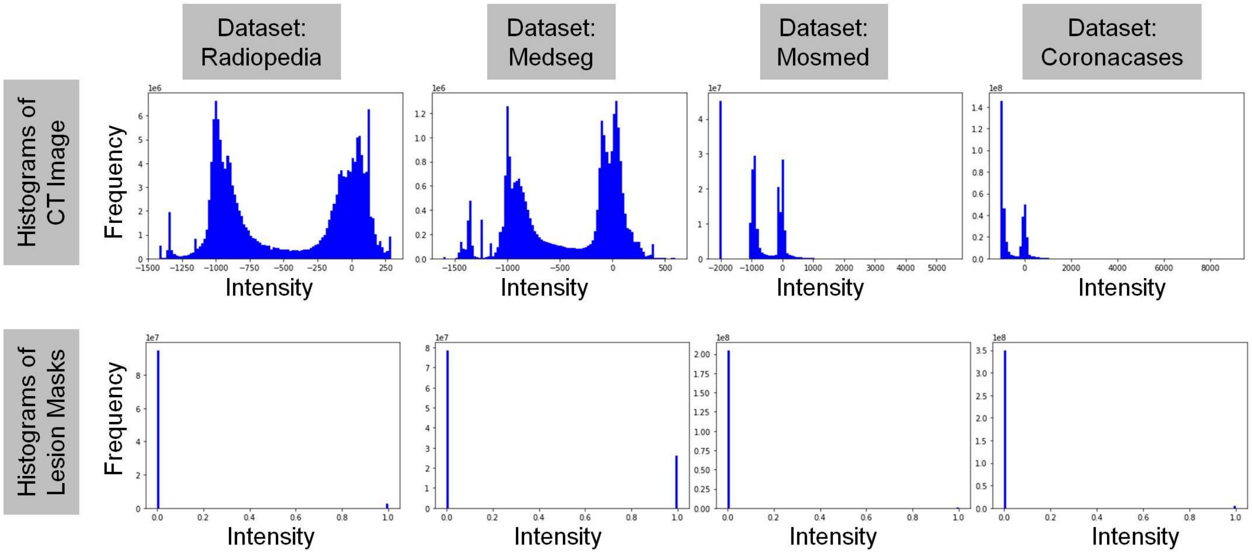


Figure S4: Representative histograms of the four CT image data-set and their masks denoting the lesion areas.

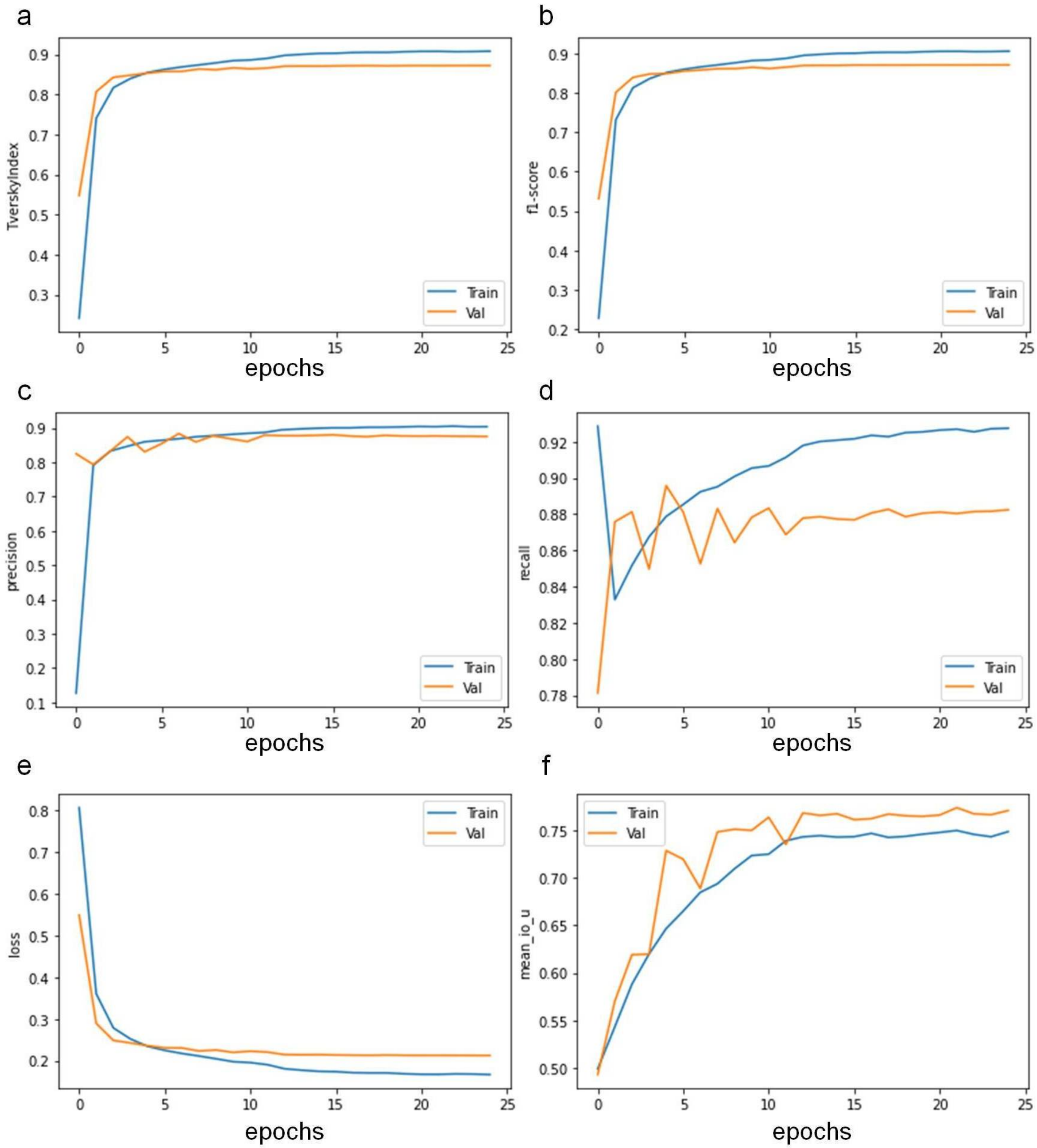


Figure S5: Performance evaluation of lesion detection by the deep learning framework showing (a) Tversky index, (b) F1-score, (c) Precision, (d) Recall, (e) Loss, and (f) Mean IoU (intersection over union).

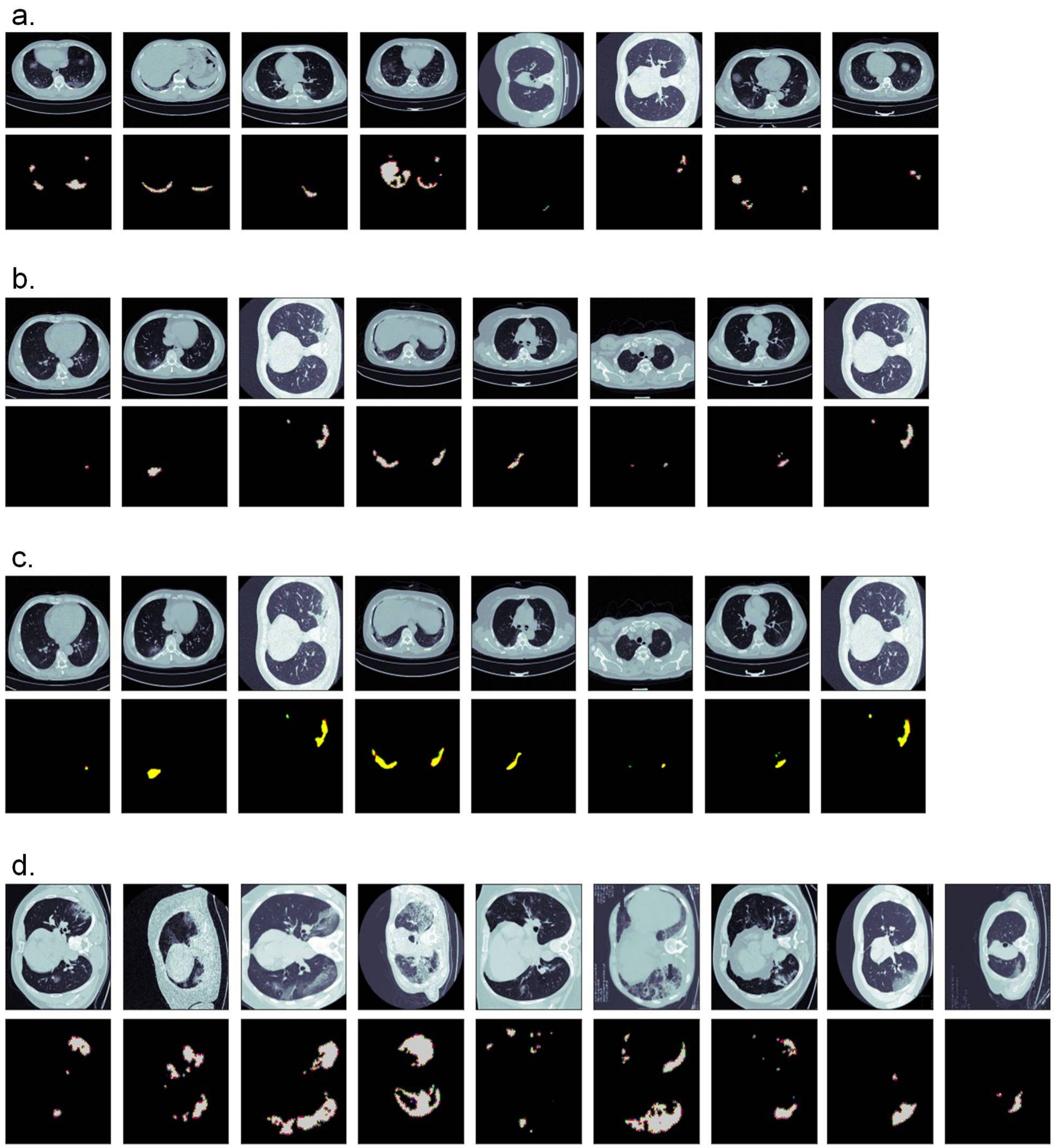


Figure S6: Representative CT images with detected lesion areas from training, validation and testing using the deep-learning framework. (a) Images from training, (b) images from validation, (c) prediction of model on validation set, and (d) prediction of model on test set. For the prediction of validation set in (c) the color code is—Yellow=true positive, Red=false-positive, Green=false-negative.

### SUPPLEMENTARY TABLES

Table S1: Patient clinical data-sheet

| **Case #** | **Age** | **Sex** | **Clinical Findings** | **Radiological Features** | **Clinical Severity** |
|---|---|---|---|---|---|
| C2 | 62 | M | pO2:97%<br>fever, cough and asthenia | scattered blurred ground-glass opacities suspected<br>for early-stage interstitial pneumonia | S1 |
| C10 | 60 | M | fever for three days | defined ground glass opacities | S1 |
| C11 | 55 | | persistant fever<br>recently underwent<br>prostatectomy | consistent with interstitial pneumonia | S1 |
| C16 | | | cough and fever for one week<br>not responding to antibiotics<br>admitted to the ED for respiratory<br>distress | consistent with interstitial pneumonia | S1 |
| C21 | 71 | M | Temp:37.8°C<br>pO2: 97% | ground-glass opacities with predominant subpleural<br>distribution in upper lobes and in the apical segment<br>of the lower lobes. | S1 |
| C28 | 48 | F | Fever<br>Blood test: leukopenia. | ground-glass opacifcations | S1 |
| C33 | 55 | M | | multiple ground-glass opacities, no pleural<br>effusion or mediastinal adenopathies. | S1 |
| C56 | 52 | F | 4 days fever<br>Normal blood count,<br>PCR 10.12 mg / L,<br>PCT 0.13 ng / mL;<br>LDH 279 U / L. | affecting the postero-basal segment of the left lower<br>lung lobe, ground-glass pseudonodular thickening of<br>the parenchyma, suspected for initial phase<br>of SARS-CoV2. Small dorsal subpleural striae<br>affecting the upper segment of the<br>right lower lung lobe. | S1 |
| C61 | 70 | M | Temp:37.8°C<br>pO2:97% | presence of multiple ëmery glass parenchymal<br>thickenings, located in the upper lobe of both lungs, in<br>the middle lobe and in the lower lobe of both lungs,<br>especially on the right, arranged sub-pleurally. | S1 |
| C62 | 56 | M | pO2:97%<br>reports fever in the last week<br>even though he has been apyretic<br>for a day, however anosmia,<br>ageusia, pharyngodynia and<br>arthralgias persist | demonstrates two ëmery glass parenchymal<br>thickenings located in correspondence of the<br>apical segment of the lower lobe of the right lung<br>subpleurally and of the mediobasal segment of<br>the lower lobe of the left lung | S1 |

| | | | | | |
|---|---|---|---|---|---|
| C63 | 49 | M | Temp:39°C<br>pO2:94%<br>fever and dyspnea for about 4 days;<br>reports hyperpyrexia | presence of multiple nuanced emery glass parenchymal thickenings located in the upper lobe of the right lung, in the middle lobe and in the lower lobe of both lungs in the subpleural area. | S1 |
| C69 | 58 | F | pO2:99%<br>Temp:36.8°C | multiple and bilateral areas of increased peri-broncovasal ground glass density, prevalent in the lower right lung lobe, suspected for localization of interstitial pneumonia; concomitant bilateral pleural effusion of modest entity on the right and minimal entity on the left, with multiple ilo-mediastinal lymphadenomegalies,<br>the major right pulmonary hilar with a maximum short axis of 13 mm. | S1 |
| C71 | 88 | M | Temp:37.1°C<br>pO2:96%<br>diabetes and high blood pressure,<br>pain in abdomen on deep palpation, positive Murphy, valid peristalsis, pure heart tones with free breaks<br>ESR: 131 mm/hr, Fibrinogen: 487mg/dl,<br>Lymphocytes: 13.1%, AST: 114UI/L,<br>ALT: 114UI / L, GAMMA GT 188 IU/L,<br>12$\mu$g / dl Sideremia,<br>Ferritin: 4239.00 ng / ml,<br>PCR: 12.20 mg / L,<br>Procalcitonin: 0.13 ng / ml.<br>Mild respiratory alkalosis<br>(pH 7.44, pCO2 34 mmHg),<br>O2 Hb: 33.4% COHb: 1.7%. | ground glass, diffuse bronchitic finding | S2 |
| C72 | 77 | M | cough and dyspnoea,<br>thoraco-abdominal pain, | ground glass opacity with peripheral distribution and associated thickening of the interlobular septa, absence of pleural effusion and absence of significant ilo-mediastinal lymphadenopathies characterize the TC pattern | S2 |

| | | | | | |
|---|---|---|---|---|---|
| C73 | 58 | M | lymphopenia, high PCR | | S2 |
| C76 | 83 | M | Temp:38.9°C<br>pO2:92%<br>arterial hypertension,<br>diabetes mellitus, IPB. | nuanced parenchymal thickening in the middle<br>and lower field in the right hemithorax<br>and in the middle field on the left, multiple<br>"frosted glass" areoles with greater peripheral<br>and parascissural distribution tending towards<br>confluence, with initial consolidation<br>phenomena, in the context of which there is a<br>thickening of the mainly intralobular interstice<br>with "crazy paving" aspects | S2 |
| C77 | 56 | M | Temp:37.1°C<br>pO2:90%<br>arterial hypertension<br>lymphopenia, high PCR | multiple pseudo-nodular thickenings of para-<br>centimetric dimensions, multiple areas of<br>parenchymal thickening with "ground glass"<br>density involving both lungs. | S2 |
| C78 | 44 | F | pO2:59%<br>dyspnea, cough and hyperpyrexia. | multiple parenchymal thickenings bilaterally,<br>with irregular margins, some with air<br>bronchogram and others with ground glass<br>associated intra- and inter-lobular septal<br>thickening, prevalently peribroncovasal and<br>subpleural distribution and with total<br>engagement of the lower lobes, in particular<br>the LID ; thin fibrotic stria in the LIS. | S2 |
| C80 | 77 | M | Temp: 37.2°C<br>pO2:89%<br>neutrophilic leukocytosis,<br>lymphopenia, modest<br>monocytopenia,<br>progressive and marked rise in PCR. | in the lower lobes, extensive ground-glass patches<br>are observed, with initial thickening of<br>the inter- and intra-lobular septa ("crazy paving"<br>pattern) and aerial bronchogram; the major,<br>subpleural, are appreciated in the dorsal sectors<br>(LIS, LID), the minor ones in the LID, in the<br>infracardiac (paravertebral and epidiaphragmatic<br>subpleural) and posterobasal (both central<br>and epidiaphragmatic subpleural) segments,<br>respectively.glass with prevalently peri-<br>broncovasal and subpleural distribution and<br>higher level in the lower lobes, particularly LID. | S2 |

| | | | | | |
|---|---|---|---|---|---|
| C82 | 49 | F | pO2:85%<br>leukopenia (especially lymphopenia);<br>increase in PCR and LDH. | presence of multiple and nuanced areas of<br>hypodiaphaly, partly pseudonodular in<br>appearance, borne by both hemithorax, with a<br>predominantly peribronchial distribution, with<br>associated reinforcement of the interstitial texture<br>and peribronchial thickening, more<br>evident in the ilo-peri-ilar and mediobasal, bilaterally.<br>In the basal center there are some<br>areas delimited by radiopaque streaks,<br>possible emphysematic manifestations.<br><br>Follow up:compared to the previous examination,<br>there is a clear reduction in the extension<br>of the previously reported densities; focal areas<br>of altered density remain with a"ground glass"<br>appearance at the level of the upper lobes and<br>thickening with a lamellar appearance in the left<br>basal area (the latter due to partial re-expansion<br>of the previous lung parenchyma involved).<br>The remaining finds are unchanged. | S2 |
| C84 | 61 | M | pO2:88%<br>fever for about 10 days,<br>cough and respiratory failure<br>a significant increase in the values<br>of procalcitonin, LDH and PCR. | widespread picture of ground glass is documented<br>in both pulmonary parenchymes with peripheral<br>distribution, mainly affecting the lingula and the<br>lower lobes (in particular in the posterior segments). | S2 |
| C85 | 47 | F | fever and dyspnea | alveolar infiltrates are recognized in the bilateral<br>intercleidoilary site and in the right middle field. No<br>pleural effusion. Cardiac transverse diameter increase | S2 |
| C89 | 44 | F | pO2:59%<br>dyspnea, cough and hyperpyrexia | Multiple parenchymal thickenings bilaterally, with<br>irregular margins, some with air bronchogram<br>and others with "ground glass associated intra- and<br>inter-lobular septal thickening, with prevalent peri-<br>broncovasal and subpleural distribution and prevalent<br>peribroncovascular and subpleural distribution and<br>with subtotal lobe engagement lower, in particular of<br>the LID; thin fibrotic stria of the LIS. | S2 |

| | | | | | |
|---|---|---|---|---|---|
| C93 | 66 | M | pO2:88%<br>reported fever (for at least 5 days)<br>and worsening dyspnea | a widespread ground glass picture is documented in<br>both lungs, mainly affecting the middle lobe, the lingula<br>and the lower lobes. | S2 |
| C96 | 45 | F | pO2:92% | bilaterally there are some patches of parenchymal<br>thickening with a "ground glass" appearance,<br>with a prevalently subpleural distribution. There are<br>multiple areas of parenchymal consolidation<br>bilaterally, particularly at the level of the dorsal<br>segment of the right upper lobe and the<br>postero-basal parenchyma of both lower lobes.<br>Absence of pleural, pericardial effusion and<br>mediastinal lymphadenopathy. The presence of<br>three solid nodular formations is also noted, the largest<br>of which (10 mm) is located in the lateral-basal segment<br>of the left lower lobe. | S2 |
| C98 | 77 | M | pO2:95%<br>ischemic heart disease,<br>high blood pressure<br>and diabetes (being treated).<br>Troponin: 0.315 ng / ml -<br>Myoglobin: 416.97 ng / ml -<br>CPK: 354 U / l;<br>Severe lymphopenia; High PCR | bilaterally, multiple and widespread parenchymal<br>thickening patches with "ground glass" appearance<br>and confluent character are observed, with mantle<br>and peribronchial distribution with associated<br>widespread thickening of the interlobular septa<br>("crazy paving pattern"). Cluster bronchiectasis is<br>appreciated in the upper lobar region on both sides,<br>particularly on the right. | S2 |
| C99 | 75 | M | pO2:94%<br>trilinear cytopenia,<br>only lymphopenia is marked<br>(540 / mmc).<br>Altered inflammation indices<br>(in particular ferritin 5723 mcg / L,<br>PCR 59 mg / L).<br>Fibrinogen 5.36 g / L.<br>Creatinine 1.65 mg / dL.<br>Mild hypoalbuminemia (32.8 g / L). | parenchymal thickening and crazy-paving patterns. | S2 |

| | | | | | |
|---|---|---|---|---|---|
| C100 | 51 | M | pO2:90% | the thickenings which previously had a "ground-glass" character, now appear to be replaced by diffuse reticular bands similar to fibrotic which are associated with thickening of the small interstitium. These findings appear ubiquitously localized, but particularly evident in the bilateral mantle with distribution tending to confluence. These also appeared in the left apical area, where previously the lung parenchyma was scarcely affected by the pathology | S2 |
| C101 | 75 | F | fever and dyspnea, diabetic increased PRC, LDH, D-dimer and IL-6 values | frosted glass thickenings with prevailing bilateral mantle arrangement, more evident on the left, where they tend to confluence. On the right, conversely, these thickening have a pseudonodular, patchy appearance. | S2 |
| C103 | 71 | M | Temp:37.8°C<br>pO2:95%<br>diabetic<br>Laboratory tests show leukopenia with neutrophilia, increase in PCR values | presence of different "ground glass" parenchymal thickenings with mainly subpleural mantle distribution, particularly evident in the postero-lower sectors of bothlungs, compatible with medium-high viral pneumonia. | S2 |
| C105 | 57 | M | suffering from fever, cough and hyperpyrexia refractory to antipyretics. | acute inflammatory lung damage in fibrotic structural modifications of the parenchyma with honey-comb like pattern. | S2 |
| C111 | 57 | M | Temp:38.5°C<br>pO2:85%<br>leukopenia; significant increase in PCR, procalcitonin and LDH values. The values of D-Dimero and Troponina were within the limits. | HRCT has documented, in both lung parenchyma, the presence of multiple thickenings with a "ground glass" appearance and some areas with a "crazy-paving" pattern, due to the coexistence of "ground-glass" areas, of interstitial consolidation and thickening. A few small reactive lymph nodes in the ilo-mediastinal area. Cardiac mage within limits. No evidence of pericardial effusion. Minimal bilateral basal pleural effusion. | S2 |

| | | | | | |
|---|---|---|---|---|---|
| C112 | 70 | M | Temp:39°C<br>pO2:80%<br>Arterial hypertension and carotid atheromasia. | chest X-ray worsened compared to the entrance, as pulmonary parenchymal thickening increased, now presenting an aspect tending to confluence, up to complete bilateral pulmonary opacification (more evident on the right). There was also bilateral interstitial involvement of a reticulo-micronodular character. Heart shadow within limits. | S2 |
| C74 | 38 | M | leukopenia (especially lymphopenia), PCR at the upper limits | ground-glass opacifications observed | S1 |
| C75 | 66 | F | pO2:92% | multiple confluent parenchymal thickenings, with ëmery glass density at the level of both lungs, with a predominantly subpleural distribution, with thickening of the inter-lobular andintra-lobular septa. | S1 |
| C91 | 78 | F | | widespread moderate interstitial peribroncovasal pulmonary reinforcement with nuanced reduction of the diaphanous to mantle site of the left upper lung field (alveolar-interstitial engagement); Deserving of diagnostic study by means of chest CT; no radiographic signs of pleural effusion and PNX bilaterally. | S1 |
| C95 | 39 | F | Hematochemical tests substantially normal, with the exception of monocytosis (monocytes 16.0%), in particular WBC and C-RP within limits. | multiple nuanced mantle areoles with ground glass in the dorsal sectors of both lower lung lobes are appreciable, without parenchymal consolidation. In the lung segments not affected by the findings described above, there are no widespread alterations of the interstitium. No pleural effusion. | S1 |
| C97 | 31 | F | Temp:39°C<br>pO2:96% | multiple areas of parenchymal thickening with a “ground-glass” appearance and some focal areas of consolidation, available bilaterally and mainly sub-mantle, in particular on the right. Autonomous origin from the trachea of the bronchi for the apical and anterior segments of the LSD. Absence of pleural, pericardial effusion and mediastinal lymphadenopathy. | S1 |
| C102 | 51 | M | fever, irritating cough, dyspnea<br>increase in PCR and LDH values | documents some circumscribed ground-glass pseudo-nodular parenchymal thickening in both lungs, with a prevalent mantle distribution, compatible with mild viral pneumonia. | S1 |

| | | | | | |
|---|---|---|---|---|---|
| C104 | 48 | F | | the examination demonstrates extensive ground-glass thickening, involving part of the anterior segment of the upper lobe and part of the medial segment of the middle lobe. On the left, only a circumscribed subpleural thickening is seen in the lateral segment of the lower lobe. The finding seems to be attributable to an atypical form of viral pneumonia. | S1 |
| C106 | 42 | M | | there are multiple areas with ground glass, partly confluent, with subpleural distribution, from the apexes to the bases, more evident on the right | S1 |
| C107 | 20 | F | pO2:99% | demonstrates the presence of three millimetric pseudo-nodular thickenings (arrows) in the posterior slopes of the lower right lobe with subpleural arrangement, which, although unspecific, are to be referred to viral pneumonia in the mild phase. Minimal thickening of the adjacent pulmonary interstitium. No parenchymal lesions on the left. Pleural effusion and mediastinal adenopathies are not appreciated. | S1 |
| C108 | 61 | F | Temp:37.3°C<br>cardiac, hypertensive<br>and diabetic patient | there are some ground-glass parenchymal thickenings with a prevalent "patchy" subpleural distribution, more numerous on the left (anterior and posterior segment of the upper lobe, lingula, lateral segment lower lobe) than on the right (lateral segment of the middle lobe and, pseudonodular, in the lateral basal segment of the lower lobe). In reconstructions with mediastinal filter various lymphadenopathies in the superior and prevascular paratracheal area are also detected, the largest of which has a short axis of about 17 mm and causes compression on the adjacent lung parenchyma (anterior segment upper left lobe, fig. 10, arrow). Other more limited lymphadenopathy is documented in the lower paracaval area. | S1 |
| C109 | 47 | M | pO2:95%<br>increase in fibronogen (511 mg / dl)<br>and ESR (23 mm / h) values | parenchymal thickening in frosted glass with associated thickening of the interlobular septa, with peripheral distribution | S1 |

| | | | | | |
|---|---|---|---|---|---|
| C110 | 39 | M | increase in transaminase values<br>and a decrease in CK-MB values<br>(0.88 ng / ml) and myoglobin<br>(23 ng / ml). | demonstrates nuanced ground-glass thickenings at the<br>anterior and posterior segments of the upper right lobe,<br>in the apicodorsal segment of the upper left lobe and in<br>the upper segment of the lower left lobe. At this level,<br>concomitant thickening of the interlobular septa is observed. | S1 |
| C115 | 51 | M | Temp:38.9°C<br>fever, chest tightness<br>and mild dyspnea<br>tests showed normal values,<br>especially white blood cells,<br>neutrophils and lymphocytes. | partial regression of the known GGO alterations<br>previously reported to the LSS and lingula | S1 |
| C81 | 74 | F | Temp:38.5°C<br>pO2:90%<br>EGA performed with PO2 of 51mmHg,<br>PCO2 36mmHg, PH 7.48 Sat O2 88%.<br>WBC 3.7 × 1000, Hb 12.6 g / dL,<br>PCR 2.22 mg / dL,<br>creatininemia 0.95mg / dL,<br>blood sugar 119mg / dL,<br>Procalcitonin 0.04 ug / L. | small ground glass areoles on the pulmonary periphery,<br>center-lobular, of phlogistic-inflammatory significance are reported | |
| C113 | 90 | F | Temp:38°C<br>pO2:88%<br>Alzheimer's disease,<br>COPD, arterial hypertension<br>and widespread multi-district<br>calcific parietal atheromasia.<br>Anemia. Increase in PCR, LDH and<br>procalcitonin values. The D-Dimer and<br>troponin values were within the limits | prominent GGOs observed | S1 |
| C114 | 20 | F | Temp:38.5°C<br>pO2:90% | in correspondence of the apico-dorsal segment of the LIS, the<br>presence of a parenchymal consolidation area with<br>subpleural distribution extending caudally to the postero-<br>basal segment, with air bronchiologram in the<br>context, which is associated with nuanced parenchymal<br>thickening with a "frosted glass" appearance.<br>Located in the adjacent seat. | S1 |

| | | | | | |
|---|---|---|---|---|---|
| C79 | 61 | M | pO2:59% | at LSD, LSS, LM and lingula multiple areas of increased pulmonary density with ground glass opacity associated with inter- and intra-lobular septal thickening, with prevalently peri-broncovasal and subpleural distribution;parenchymal thickening, with patent air-bronchogram, in the lower lobes. | S3 |
| C86 | 78 | M | Temp:38.2°C<br>pO2:92% | with confirmation of widespread and bilateral foci of parenchymal consolidation, with relative saving of the apexes, which are associated with areas with a ground glass appearance; coexists modest bilateral pleural effusion, of greater entity on the left: compatible with interstitial pneumonia with consolidative aspects. No filling defects of a thrombo-embolic nature are observed within the large branches or main branches of the pulmonary artery. | S3 |
| C90 | 61 | M | Temp:38.2°C<br>pO2:92% | in LSD, LSS, LM and lingula plurime areas of increased lung density ”with frosted glass which is associated with inter- and intra-lobular septal thickening, with a prevalent peribroncovasal and subpleural distribution; parenchymal thickening, with patent air bronchogram, to the lower lobes. | S3 |
| C94 | 64 | F | elevated fever,<br>persistent cough and dyspnoea | bilateral parenchymal thickening vidence of bilateral involvement with ”frosted glass” areas, | S3 |
| C3 | 57 | M | hypoxemia and hypocapnia | subpleural ground-glass opacity with predominant subpleural distribution and consolidations in all lobes, particularly in the lower lobes | S3 |
| C5 | 63 | M | | ground-glass opacities in the lower lobes are more extended and consolidations are also noted. | S3 |
| C6 | 32 | F | treatment with steroids<br>for autoimmune disease<br>Fever and cough, leukocytosis<br>(hypoxemia and hypocapnia | diffuse bilateral consolidations partially sparing the upper lobes and the apical segments of the lower lobes. | S3 |
| C9 | 73 | F | | diffuse ground-glass opacity with confluent consolidations in the dependent areas and peripheral band atelectasis | S3 |
| C17 | 80 | M | cardiac failure<br>fever, dyspnea and cough | confluent consolidations | S3 |
| C20 | 75 | M | C-RP 15mg/dl, Procalcitonin: 8.9mg/dl | multiple and predominantly subpleural “ground-glass” opacities with reticulation and consolidations, involving all lobes, especially the upper lobes. | S3 |

| | | | | | |
|---|---|---|---|---|---|
| C24 | 78 | F | pO2:50%<br>asthenia, sick cough and fever<br>for 3 days | diffuse ground-glass opacities with reticulation in a "crazy-<br>paving" pattern, associated with alveolar consolidations<br>in the dependent regions. | S3 |
| C25 | 71 | F | COPD, Diabetes mellitus,<br>chronic renal failure, mitral<br>valve replacement | multiple and large ground glass with reticulations with<br>"crazy-paving" pattern associated with consolidations.<br>Right pleural effusion. | S3 |
| C38 | 72 | F | Temp:36.9°<br>pO2:92%<br>diabetic | bilateral parenchymal consolidations in the posterior regions<br>of the lower lung lobes, bilaterally. Subpleural<br>nodules at the anterior segments of the left upper lung lobe.<br>Multiple ground glass opacities over the entire lung area | S3 |
| C41 | 73 | M | | bilateral confluent consolidations prevalent in the upper<br>lung fields associated with ground glass areas configuring<br>a widespread picture of "crazy paving" with relative saving<br>of the subpleural regions. Bilateral parenchymal bands | S3 |
| C42 | 87 | M | pO2:95%<br>diagnosis of right heart failure<br>in hypertensive heart disease<br>and PM, regressed with diuretic<br>therapy | large areas of ground-glass interstitial thickening with<br>initial areas of larger consolidation in the posterior basal<br>segments, in the absence of pleural effusion | S3 |
| C44 | 65 | M | Temp:37.5°C<br>pO2: 84% | wide and bilateral sloping and symmetrical "ground glass"<br>aspect, with aerial bronchogram and moderate tendency<br>to form small consolidations in the posterior subpleural<br>area. By sparing only the apices, the alterations extend<br>to the bases and have an antero-posterior thickness of<br>6-9 cm. Cortical regions are not spared. There is no pleural<br>or pericardial effusion and the interlobular septa are not<br>significantly thickened-imbibed. An emphysematous bulla<br>at the left base of the lung in the posterior median area of 2.7 cm | S3 |
| C45 | 79 | | fever and cough<br>diabetic, cardiopathic | bilateral interstitial pneumonia. | S3 |
| C46 | 61 | M | Temp:38.5°C<br>pO2:85% | bilateral mantle consolidations with air bronchogram affect the<br>lower lobes of both lungs, especially the apical segments,<br>and the dorsal and apico-dorsal segments of the upper<br>lobes of the right and left respectively. The remaining<br>segments of the upper lobes present thickened areas with<br>ground glass opacity. | S3 |

| | | | | | |
|---|---|---|---|---|---|
| C47 | 61 | M | pO2:82%<br>WBC 11.2; PCR 301 mg / L<br>(VN <5);<br>LDH 738mg / dl<br>(VN 135-225);<br>Fibrinogen 798mg / dl | multiple areas of ground-glass extended diffusely in<br>both lungs with initial consolidation aspects in the basal<br>regions where subpleural sparing is highlighted; moderate<br>volume reduction of both lungs. | S3 |
| C51 | 55 | F | pO2:50%<br>dyspnea, fever and cough<br>High PCR; Procalcitonin and<br>normal laboratory tests | multiple parenchymal thickenings of the pseudonodular<br>type with density ”ground glass” at the upper lobes,<br>with a predominantly peribroncovasal and subpleural distribution,<br>with associated areas of parenchymal thickening with an air<br>bronchogram at the LID and<br>LIS. | S3 |
| C52 | 57 | M | pO2:50%<br>acute dyspnea and therapy-<br>resistant hypepyrexia | multiple areas of increased “frosted glass” lung density,<br>in particular in the upper and middle lobes, with a prevalent<br>peribroncovasal and subpleural distribution, which is<br>associated with inter- and intra-lobular septal thickening;<br>areas of parenchymal thickening, with patent air<br>bronchogram in context, at the level of the lower<br>lobes. | S3 |
| C58 | 60 | M | Dyspnea and hyperthermia<br>APR: DMT2, dyslipidemia,<br>high blood pressure,<br>ex-heavy smoker | multiple “frosted glass” thickening areas, prevalent in<br>the lower lobes and in the posterior<br>seat, with iteress of the intralobular interstitium,<br>findings compatible with acute inflammation | S3 |
| C65 | 74 | M | pO2:94%<br>history of ischemic heart<br>disease, bronchial asthma<br>and NAO therapy. | pattern characterized by the presence of “emery glass”<br>areas superimposed on smooth thickening of the interlobular<br>and intralobular interstitium; concomitant area of parenchymal<br>consolidation in the posterior cavity of the right hemithorax<br>and fibrotic outcomes in the posterior cavity of the left hemithorax | S3 |
| C67 | 83 | M | Temp:38.9°C<br>pO2:92%<br>ex-smoker with a history<br>of remote pathology positive<br>for arterial hypertension,<br>diabetes mellitus, IPB.<br>increase in PCR,<br>LDH and transaminases | multiple ”frosted glass” areoles with greater peripheral and<br>parascissural distribution tending to confluence, with initial<br>consolidation phenomena, in the context of which there is a<br>thickening of the intralobular interstice with ”crazy<br>paving” aspects. | S3 |

| | | | | | |
|---|---|---|---|---|---|
| C1 | 80 | M | fever and dyspnea | multiple ground-glass opacities associated with reticulations in a “crazy-paving pattern” – particularly in the lower lobes. Subpleural consolidation | S2 |
| C4 | 45 | M | fever, cough | multiple diffuse ground-glass opacities in all lobes, with random distribution, predominant subpleural and peri-bronchial in the upper lobes | S2 |
| C7 | 43 | M | Respiratory failure (hypoxemia and hypercapnia<br>Leukopenia | subpleural ground-glass opacity with consolidations in the LLL; scattered ground-glass opacities in the upper lobes and more extended in the lower lobes | S2 |
| C14 | 50 | M | pO2:93%<br>sick cough asthenia and fever for two days | crazy paving patterns observed | S2 |
| C15 | 46 | F | pO2:98% | Multiple and bilateral consolidations at different stages, with predominant peribronchial and subpleural distribution. No pleural effusion | S2 |
| C18 | | | Cough, fever and progressive dyspnea<br>Colon cancer | ground glass opacifications and septal thickening | S2 |
| C19 | | | Temp:39°C<br>Acute sinusitis fever and cough. Onset of asthenia, diarrhea and hyporexia since the day before the admission to the ED. | ground glass opacifications and septal thickening | S2 |
| C22 | 69 | F | Temp:36.9°C<br>pO2:96% | multiple ground-glass opacities with predominant sub-pleura distribution associated with areas of “crazy-paving” pattern. | S2 |
| C23 | 27 | M | pO2:92 | multiple ground-glass opacities and alveolar consolidations with predominant subpleural distribution in the upper and lower lobes. | S2 |
| C27 | 58 | F | dyspnea and fever | interstitial markings with alveolar consolidations in the upper lobes. Cardiomegaly. No pleural effusion | S2 |
| C29 | 68 | M | fever, dyspnea and diarrhea<br>eukocytosis, increased C-PR, procalcitonin in the range<br>chronic lymphocytic leukemia under follow-up, Dyslipidemia | multiple and bilateral scatterd ground-glass opacities with predominant subpleural distribution associated with reticulations and alveolar consolidations | S2 |

| | | | | | |
|---|---|---|---|---|---|
| C30 | 64 | M | dyspnea, cough and fever<br>C-PR 13.44 mg/dl,<br>procalcitonin and CBC<br>unremarkable.<br>Diabetes mellitus | bilateral scatterd ground-glass opacities with<br>predominant subpleural distribution | S2 |
| C31 | 63 | M | fever, pharyngalgia asthenia and<br>diarrhoea<br>C-PR 16.27 mg/dl,<br>procalcitonin and CBC<br>unremarkable.<br>Obstructive sleep apnea<br>syndrome | multiple and bilateral ground-glass opacities<br>with consolidations | S2 |
| C32 | 43 | M | fever and asthenia | large ground-glass opacity with reticulations<br>and small peripheral consolidations in<br>the RLL; scattered GGOs in the LUL. | S2 |
| C34 | 47 | M | Temp:39°C<br>pO2:95% | development of a dense consolidative pattern<br>and interstitial bands, consistent with<br>good evolution of the disease. | S2 |
| C35 | 63 | | leukopenia, an increase in<br>transaminases | interstitial-alveolar pneumonia with widespread<br>bilateral mosaic alteration with crazy paving pattern. | S2 |
| C36 | 76 | M | | multiple Pulmonary thickenings, mainly of the<br>"ground glass" type with ubiquitous bilateral<br>distribution. Air flap in the peritoneal cavity<br>compatible with intestinal perforation and free<br>intra-abdominal effusion. | S2 |
| C37 | 86 | M | dyspnea, fever and cough | "frosted glass" type thickening with bilateral<br>subpleural distribution. Concomitant thickening of<br>the interlobular septa (crazy paving pattern). | S2 |

| | | | | | |
|---|---|---|---|---|---|
| C40 | 56 | | Temp:38.5 slight reduction in white blood cells (3.47 $x10^3$ / ul; vn 4.5 -10.0) in the absence of significant lymphopenia,increased fibrinogen (650 mg / dl; vn 150-450 ), negative troponin, ESR within the limits ( 13 mm / h; vn <15). | the CT examination documents in both lung areas the presence of multiple areas of increased and altered parenchymal density with ”frosted glass tending to confluence with consolidation, of which the largest at the apical segment of the right upper lobe, apico-dorsal lobe upper left, lower lingular and more extended to the basal pyramids on both sides, with predominantly mantle arrangement. No signs of pleural or pericardial effusion. Regular patency of the trunk and main pulmonary arteries. Diffuse sub-and peri-centimetric reactive lymphadenopathy in the peritracheo-broncho-hilar chains on both sides. | S2 |
| C43 | 82 | F | WBC 6.84; PCR 106.93 mg / L (limit 5); VES 45 (limit 15); LDH 314 mg / dl (limit 214); Glucose 137mg / dl | Slight hypo-expansion of the left lung in which context multiple areas of ground glass are recognized with associated smooth thickening of the inter- and intra-lobular interstitium. The alterations are prevalent in the subpleural interstitium and mainly localized to the dorsal and lateral sectors of the upper left lobe; on the lower left lobe some consolidated supbleuric aspects are found | S2 |
| C49 | 84 | M | High fever, recent onset of moderate respiratory failure since 2 weeks. | widespread ”crazy paving areas of ”ground-glass” with aspects of confluence at the bases. Thin bilateral pleural effusion and small sub-centimetric mediastinal oval lymph nodes are associated, | S2 |
| C50 | 60 | M | abdominal pain and hyperpyrexia lymphocytes 1.1 x $10^9$ / L | | S2 |
| C53 | 78 | | pO2:84% fever and cough for one week and respiratory failure | multiple frosted glass opacities affecting all the lobes to a greater extent the upper lobes and the lower left lobe m, in the context of which there is widespread thickening of the interstitial septa. Mediastinal adenopathies. | S2 |

| | | | | | |
|---|---|---|---|---|---|
| C54 | | M | arterial hypertension | frosted glass thickenings with peripheral distribution and associated thickening of the interlobular septa, absence of pleural effusion and in the absence of significant ilo-mediastinal lymphadenopathies characterize the TC pattern | S2 |
| C55 | 65 | F | bariatric surgery, bipolar syndrome | all lung segments are affected by numerous patches of parenchymal thickening with emery glass density, some with confluent appearance, without pleural effusion or signs of pulmonary thrombo-embolism | S2 |
| C57 | 40 | M | Temp:40°C<br>Previous pneumonia,<br>former smoker.<br>fever and from now dyspnea hypoxemia (PaO2 63.4 mmHg), mild respiratory alkalosis (pH 7.5, pCO2 35 mmHg) and to blood tests increase in inflammation indexes with: PCR: 87.16 mg / L,<br>Fibrinogen: 621 mg / dL,<br>Procalcitonin: 0.16 ng / ml,<br>LDH: 328 U / L | confirms multiple thickenings largely with a ground glass appearance, arranged in patches and involving almost all the lung lobes, with ilo-parailary and partly sub-pleural distribution: strongly suspected finding for COVID 19 pulmonary infection.<br>No pleural effusion is documented.<br>The trachea and the main bronchi remain patent.<br>On the right, in the anterior territory of the superior lobe there is a voluminous pulmonary cyst devoid of inclusions with an axial diameter of about 8 cm and craniocaudal of 10 cm. | S2 |
| C64 | | | Temp:39°C<br>pO2:90% | the image demonstrates the presence of multiple parenchymal thickenings with emery glass and some areas with "crazy-paving" patterns characterized by the presence of superimposed emery glass" areas with smooth thickening of the interlobular and intralobular interstitium with associated areas of parenchymal consolidation | S2 |
| C66 | | | Temp:39°C<br>pO2:93% | the CT survey demonstrates some small nuanced parenchymal "ëmery glass" thickenings located at the apicodorsal segment of the upper lobe of the left lung and in the middle lobe.<br>Fair share of pleural effusion in the large cavity on the right with a thin layer of effusion also on the left. Widespread manifestations of centrilobular and subpleural paraseptal emphysema in the lung parenchyma. | S2 |

Table S2: List of CT imaging centers and corresponding case numbers in the dataset

| # | **Hospital/CT Imaging Centers** | **Case Numbers** |
|---|---|---|
| 1 | U.O.C. Diagnostica per immagini – Ospedale “Dono Svizzero” Formia DEA I livello– Asl Latina | 21,22,23,24,25,27,61,62,63,64,65 |
| 2 | Radiologia ASST Cremona | 3,4,5,6,7 |
| 3 | UOC di Radiologia Diagnostica e Interventistica, Ospedale Madonna delle Grazie, ASL 4 – Matera | 10,16,53 |
| 4 | UOC Radiologia ASST Bergamo Est | 2,9,14,15,33 |
| 5 | SS Oncological Radiology San Giuseppe Moscati Hospital, Taranto | 51,52,78,79,89,90,91,115 |
| 6 | Radiologia IRCCS Reggio Emilia | 29,30,31,32 |
| 7 | ASST Pavia, hospital of Vigevano | 41,43,44,95 |
| 8 | SC Radiodiagnostics - AO “S. Croce e Carle”- Cuneo | 42,56,57,86 |
| 9 | Ospedaliera San Camillo-Forlanini, Roma | 17,49 |
| 10 | General and breast diagnosis of the West - Sanremo Hospital | 36,37 |
| 11 | Fondazione Poliambulanza Istituto Ospedaliero, Brescia | 18,19,85 |
| 12 | UOC Diagnostica per Immagini, AO Riuniti Marche Nord | 20 |
| 13 | Radiology - IRCSS Sacro Cuore Don Calabria Hospital - Veneto Region - Negrar (VR) | 38,50 |
| 14 | UOC Radiology, San Paolo Hospital, Savona | 45 |
| 15 | Diagnostics for images, Aorn SG Moscati, Avellino | 46 |
| 16 | AOU Careggi, Florence | 47 |
| 17 | AOU San Luigi Gonzaga, Orbassano | 58 |
| 18 | SOC Radiodiagnostics San Jacopo Pistoia | 67 |
| 19 | UOC Radiologia – AO Sant’Orsola – Bologna | 11 |
| 20 | Filippo Barbiera – ASP Agrigento Presidio Ospedaliero di Sciacca (AG) | 28 |
| 21 | U.O di Radiologia ASST CREMA – Ospedale Maggiore di Crema | 33 |
| 22 | Sergio MargariASST Fatebenefratelli Sacco - Milan | 69 |
| 23 | Dipartimento di Scienze Radiologiche – Scuola di Specializzazione in Radiologia | 1 |
| 24 | ASL Verbano-Cusio-Ossola | 35 |
| 25 | AORN Äntonio Cardarelli” - Naples | 40 |
| 26 | Radiology AOU Policlinico Umberto I - Sapienza University of Rome | 54,77,82 |
| 27 | UOC of Diagnostic and Interventional Radiology, Carlo Poma ASST Mantua Hospital | 55 |
| 28 | Milano, Radiologia Fatebenefratelli | 70 |
| 29 | UOC di Radiologia: Direttore: Dott. F. Pinto P.O. “Anastasia Guerriero” – Marcianise, ASL Caserta. | 71 |
| 30 | Radiologia – AOU Policlinico Umberto I – Sapienza Università di Roma | 72,73,74,75 |
| 31 | 1. SOC Radiodiagnostica Ospedale San Jacopo Pistoia<br>2. Reparto Malattie Infettive Ospedale San Jacopo Pistoia | 76 |
| 32 | USC Radiologia ASST Lodi – Presidi Ospedalieri di Codogno e Casalpusterlengo | 80 |
| 33 | Reparto di Diagnostica per Immagini Ospedale Regionale “Piccole Figlie Hospital” Parma | 81 |
| 34 | UOC Radiologia Pediatria PO G. Di Cristina ARNAS Civico Palermo | 83 |

| | | |
|---|---|---|
| 35 | Dipartimento dei Servizi Sanitari – U.O. Radiologia – P.O. “G. Jazzolino”, Vibo Valentia – ASP Vibo Valentia | 84,93,100,101,102,103,104<br>107,108,109,110 |
| 36 | 1. SOC Radiodiagnostica Ospedale San Jacopo Pistoia ASL Toscana Centro.<br>2. Reparto Malattie Infettive Ospedale San Jacopo Pistoia ASL Toscana Centro | 94 |
| 37 | “Pineta Grande Hospital” – Castel Volturno (CE), Dipartimento di Diagnostica per Immagini | 96,97,98 |
| 38 | Policlinico G.B. Rossi “Borgo Roma” – Università degli Studi di Verona – Verona (VR) | 99 |
| 39 | Dipartimento di Radiologia Asl VCO | 105 |
| 40 | Dipartimento di Radiodiagnostica e Radiologia Interventistica, Direttore D. MessanaARNAS,<br>Ospedali Civico, Di Cristina e Benfratelli, Palermo | 106 |
| 41 | Dipartimento Tecnologie Avanzate Diagnostiche e Terapeutiche,<br>U.O.C. di Radiologia – Ospedale Riuniti,<br>Azienda Ospedaliera Grande Ospedale Metropolitano (G.O.M.) “Bianchi-Melacrino-Morelli”, Reggio Calabria | 111,112,113,114 |
